# Targeting type I regulatory T cells to enhance anti-parasitic T helper 1 and T follicular helper cell responses in human malaria

**DOI:** 10.1101/2025.04.09.25325416

**Authors:** Luzia Bukali, Jessica A. Engel, Damian Oyong, Fabian de Labastida Rivera, Megan Soon, Dean Andrew, Nicholas L. Dooley, Mayimuna Nalubega, Reena Mukhiya, Teija M. Frame, Julianne Hamelink, Jessica R. Loughland, Rebecca Webster, Fiona Amante, Timothy N.C. Wells, Stephan Chalon, Joerg J. Moehrle, James S. McCarthy, Michael D. Leipold, Gerlinde Obermoser, Holden T. Maecker, Bridget E Barber, Michelle J. Boyle, Christian R. Engwerda

## Abstract

Type I interferon (IFN) signalling promotes development of type 1 regulatory (Tr1) CD4^+^ T cells, which suppress inflammation but may limit protective immunity in malaria by constraining effector Th1 cells and antibody-promoting T follicular helper (Tfh) cells. We tested whether transiently blocking IFN-driven JAK/STAT signalling with ruxolitinib would reduce Tr1 cell development and enhance protective Th1 and Tfh cell responses. Using controlled human malaria infection, we investigated the impact of ruxolitinib on CD4^+^ T cell activation and antigen-specific immunity. Whole blood stimulation with IFNβ, IL-2, or PMA/Ionomycin revealed that ruxolitinib suppressed pSTAT1, pSTAT3, and pSTAT5 in CD4^+^ T cells following treatment. However, antigen-specific CD4^+^ T cell responses were preserved. Further, ruxolitinib enhanced recall CD4^+^ T cell responses during a second infection, with increased frequencies of antigen-specific Th1, Tfh, and Tr1 cell subsets. Transcriptional analysis revealed overlapping gene signatures and clonal sharing between Th1, Tfh and Tr1 cells. These findings show transient JAK inhibition can modulate development of Th1, Tfh and Tr1 cell subsets in malaria, suppressing inhibitory signalling while enhancing the magnitude and durability of the CD4^+^ T cell recall response following a second infection. Targeting IFN-driven Tr1 cell activity therefore represents a promising host-directed strategy to improve malaria immunity.

Type I interferon (IFN) signalling promotes type 1 regulatory (Tr1) CD4^+^ T cells, which suppress inflammation but may also limit protective Th1 and T follicular helper (Tfh) cell responses in malaria. We tested whether transient blockade of IFN-driven JAK/STAT signalling with ruxolitinib would reduce Tr1 cell activity and enhance protective CD4^+^ T cell responses. In controlled human malaria infection, we assessed the impact of ruxolitinib on CD4^+^ T cell activation and antigen-specific responses. Ruxolitinib suppressed pSTAT1, pSTAT3, and pSTAT5 in CD4^+^ T cells during treatment but preserved antigen-specific responses. Notably, ruxolitinib treatment in first infection enhanced recall CD4^+^ T cell responses during second infection, with increased frequencies of antigen-specific Th1, Tfh, and Tr1 cell subsets. These findings indicate that transient JAK inhibition reshapes the balance of Th1, Tfh and Tr1 cell subsets, and enhances the magnitude and durability of the CD4^+^ T cell recall response following a second infection.

**Summary:** Blocking type I IFN signalling with ruxolitinib transiently reduces Tr1 cell activity while enhancing Th1 and Tfh cell recall responses, reshaping anti-parasitic CD4^+^ T cell-mediated immune responses in malaria.

## Introduction

Malaria remains a major global health threat, with 282 million cases and 610,000 deaths reported by the World Health Organisation in 2024. In addition to this health burden, malaria imposes significant socioeconomic costs on endemic communities (WHO, 2025). The recent approval of the pre-erythrocytic vaccines RTS,S and R21 marks a significant milestone in the development of tools to control malaria. However, protection is limited to young children, wanes over time, and falls short of the World Health Organisation’s 75% efficacy target (Olotu et al., 2016). Suboptimal vaccine performance has been linked to baseline immune characteristics (Moncunill et al., 2022; Oyong et al., 2023), which not only shape responses to re-infection and vaccination, but may also influence the efficacy of anti-parasitic drugs (Rogerson et al., 2010). A deeper understanding of these immune networks may lead to the development of approaches to modulate immunity and therefore improve vaccine efficacy (Boyle et al., 2024; Montes de Oca et al., 2016a; Nahrendorf et al., 2021).

CD4^+^ T cells are central to malaria immunity, coordinating responses against infection and regulating inflammation (Sallusto, 2016). IFNγ-producing Th1 cells promote antigen presentation by dendritic cells and activate macrophages to eliminate infected red blood cells (Tubo and Jenkins, 2014). There is significant heterogeneity in the molecules expressed by Th1 cells, in part due to tissue-specific adaptions in response to infection-mediated changes to inflammation and metabolism (Tuzlak et al., 2021). However, the inflammatory activity of Th1 cells must be counterbalanced to avoid immunopathology. In malaria, type 1 regulatory (Tr1) cells are important for this process via the secretion of IL-10 and expression of other inhibitory mediators, including co-inhibitory receptors (Boyle et al., 2015a; Boyle et al., 2017; Jagannathan et al., 2014; Montes de Oca et al., 2016c; Walther et al., 2009). While Tr1 cells limit immunopathology and thus disease (Edwards et al., 2023; Roncarolo et al., 2014; Roncarolo et al., 2018), they can also suppress Th1 and T-follicular helper (Tfh) cell development and function, potentially undermining protective immunity, vaccine efficacy, and memory (Montes de Oca et al., 2016b; Montes de Oca et al., 2016c; Zander et al., 2016). Tfh cells are essential for parasite-specific antibody responses. They support germinal centre B cell expansion and differentiation, thus driving the production of protective antibodies (Obeng-Adjei et al., 2015; Ryg-Cornejo et al., 2016). Functionally distinct Tfh subsets emerge during infection, and evidence from controlled human malaria infection (CHMI), field studies, and vaccine trials shows that Tfh2 and some Tfh1 cell subsets are particularly important for generating effective antibody responses (Bentebibel et al., 2013; Chan et al., 2022; Chan et al., 2020; Morita et al., 2011; Nielsen et al., 2021; Soon et al., 2025). Thus, the balance between Th1, Tfh, and Tr1 cell subsets is central to shaping the quality of anti-parasitic immunity in malaria.

Recent work highlights type I interferons (IFNs) as key regulators of CD4^+^ T cell subset balance. In experimental malaria models, type I IFNs suppress Th1 and Tfh cell differentiation while promoting Tr1 cell development (Nahrendorf et al., 2021; Zander et al., 2016). In CHMI studies, type I IFNs limited antigen-specific IFNγ production while enhancing parasite-specific IL-10 (Montes de Oca et al., 2016c). Additionally, STING-driven type I IFN signalling in CD4^+^ T cells directly promotes Tr1 cell differentiation in humans (Wang et al., 2023). Targeting this pathway with the JAK1/2 inhibitor ruxolitinib, co-administered with anti-parasitic treatment, boosted anti-parasitic Th1 cell responses in a pre-clinical model of visceral leishmaniasis (VL), and *ex vivo* in blood samples from VL patients (Kumar et al., 2020). We recently reported that ruxolitinib when co-administered with standard antimalarial treatment was well tolerated in healthy volunteers experimentally infected with blood-stage *Plasmodium falciparum,* attenuated the post-treatment inflammatory response, and remodelled the systemic immune response to a second infection (Chughlay et al., 2022; Webster et al., 2025). Here, in this same study cohort, we investigate the impact of ruxolitinib as a host-directed treatment on CD4^+^ T cell responses during malaria. We assess how transient blockade of type I IFN signalling can modulate the Th1, Tfh and Tr1 cell development and expansion with the goal of improving protective, anti-parasitic immunity.

## Results

### Ruxolitinib transiently inhibits JAK/STAT signalling in CD4^+^ T cells during malaria

To investigate the impact of ruxolitinib host-directed therapy on the immune response during malaria, we used samples collected from a Phase 1b clinical trial (ACTRN12621000866808) (Webster et al., 2025). In this study, 20 malaria-naïve volunteers were infected with *P. falciparum*-parasitised red blood cells (pRBCs). On day 8 or 9 post-infection, participants were randomised to receive standard anti-malarial treatment (artemether-lumefantrine) with either ruxolitinib (20mg) or placebo, all administered twice daily for three days. Fifteen participants were re-inoculated three months later (Fig. 1A) and the resultant infection treated with artemether-lumefantrine. Trial safety, impact on clinical parameters and plasma cytokine responses have recently been reported (Webster et al., 2025). To examine the impact of ruxolitinib on CD4^+^ T cell signalling, whole blood was stimulated with IFNβ, IL-2, PMA/ionomycin, or left unstimulated. Phosphoproteins were analysed at inoculation (I), treatment day (T), 1.5 days after treatment (T+1.5; after three doses of ruxolitinib or placebo), and five days after treatment commenced (T+5). Sampling strategy, immune cell subset identification, and responses to IL-6 stimulation were previously described (Webster et al., 2025).

**Figure 1.**
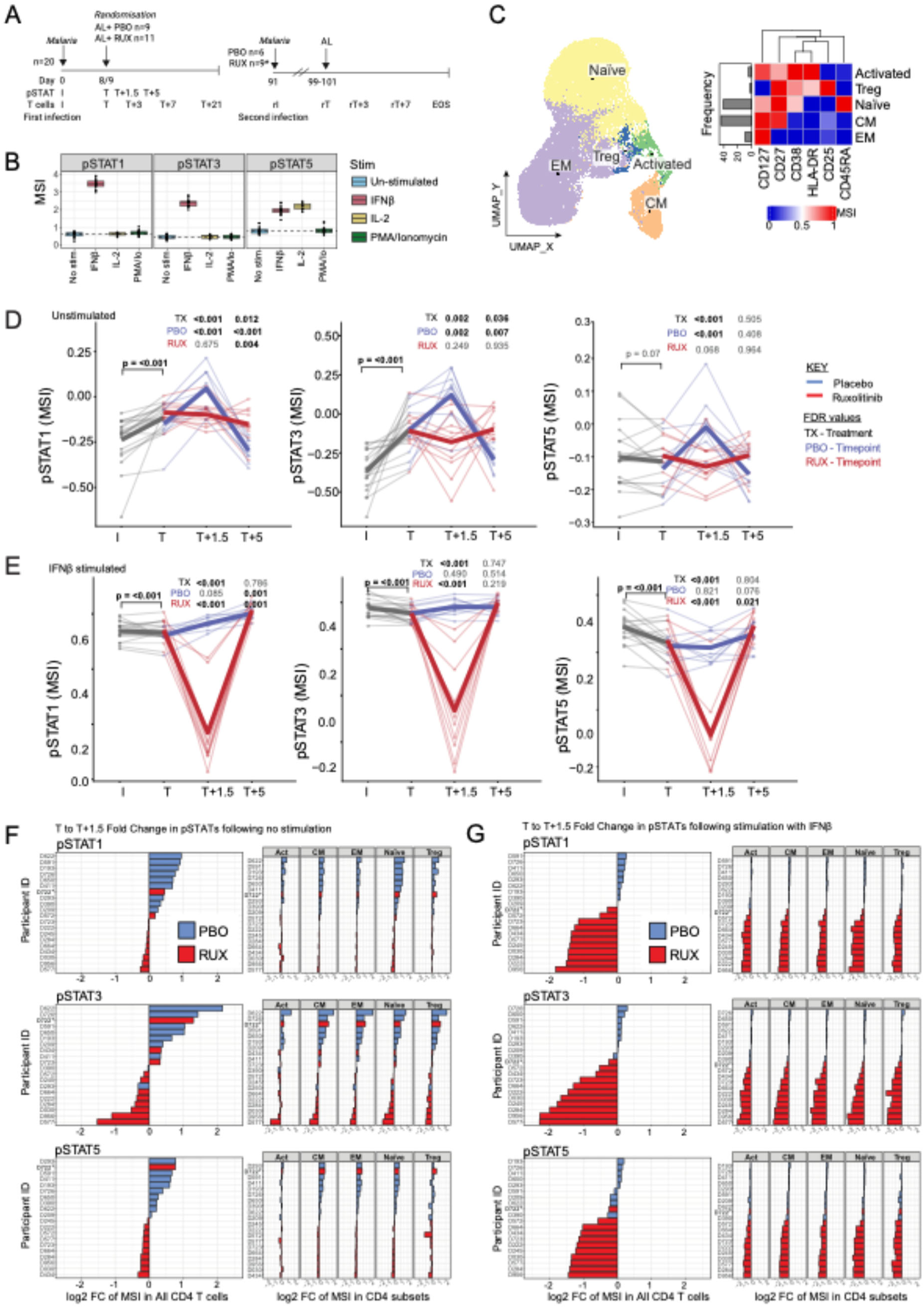
Ruxolitinib transiently reduces JAK/STAT signalling in CD4 T cells during malaria. **A)** Study design and whole blood sampling for pCyTOF of phosphoproteins (pSTAT), and T cell analysis from peripheral blood mononuclear for CyTOF, flow cytometry, activation induced marker (AIM) assays and single cell RNA and T cell receptor (TCR) sequencing (scRNA/TCRseq). Inoculation (I), re-inoculation at days 90 post-infection (rI), treatment during first (T) and second (rT) infection, as well as the end of study (EOS; ∼28 days after rT) are indicated. 20 participants were included in the first infection, and 15 in the second. * One participant in the ruxolitinib group was treated early after inoculation due to COVID-19 infection. **B)** Phosphorylation of signalling molecules during malaria in unstimulated and IFNβ, IL2 and PMA/ionomycin stimulated cells was analysed by pCyTOF at day of inoculation, treatment (T), 1.5 days after treatment (T+1.5) and 5 days after treatment (T+5). At inoculation, levels of phosphorylated (p)STAT1, pSTAT3 and pSTAT5 in CD4 T cells in unstimulated and stimulated conditions are shown. **C)** CD4 T cells were characterised by expression of CD127, CD27, CD38, HLA-DR, CD25 and CD45RA. Identified subsets and expression of markers is shown. **D/E)** pSTAT1, pSTAT3, and pSTAT5 expression in unstimulated **(D)** and IFNβ−stimulated **(E)** CD4 T cells at I, T, T+1, T+5. Data are log transformed median signalling intensities, with thin lines representing each participant and coloured by treatment group. All datapoints prior to randomisation are in grey; following randomisation ruxolitinib-treated participants are shown in red and placebo treated participants are in blue. P values are from linear mixed effect models, with bold lines representing the mean of the predicted values from the fitted models for each group. TX indicates the p values for the interaction term between each timepoint (compared to T) and treatment groups. P values for comparisons between each timepoint and T are shown for the placebo (PBO) and ruxolitinib (RUX) groups and were determined from contrasts. **F/G** Fold change in expression of pSTAT1, pSTAT3, pSTAT5 between T and T+1.5 in total CD4 T cells and CD4 T cell subsets for unstimulated (**F)** and IFNβ stimulated **(G)** cells. Data for each participant are shown. See also Supplementary Figures 1-3.

Type I interferon signalling is critically dependent on the phosphorylation of STAT transcription factors, which enables their dimerization, nuclear translocation, and induction of interferon-stimulated genes (Platanias, 2005). Prior to inoculation, IFNβ stimulation increased pSTAT1 and pSTAT3, while both IFNβ and IL-2 increased pSTAT5 in CD4^+^ T cells (Fig. 1B). PMA/ionomycin induced pCREB, pERK, and pS6 (Supplementary Fig. 1A). Based on these findings, in subsequent analyses we focused on all signalling molecules in unstimulated cells (to assess *in vivo* responses) and on molecules upregulated by stimulation (pSTAT1, 3 and 5 for IFNβ, pSTAT5 for IL-2, and pERK/pCREB/pS6 for PMA/ionomycin). CD4^+^ T cell subsets were defined by CD127, CD27, CD38, HLA-DR, CD25, and CD45RA to identify naïve, regulatory T (Treg), central memory (CM), effector memory (EM) and activated (Act) cells (Fig. 1C). Phosphoprotein expression was analysed in CD4^+^ T cells using linear mixed-effects models, comparing I to T (infection response) and T to post-treatment (treatment response, including ruxolitinib effects).

Infection upregulated pSTAT1 and pSTAT3 across all CD4^+^ T cell subsets, revealing the activation of these signalling pathways in CD4^+^ T cells during infection (Fig. 1D, Supplementary Fig. 1B). pSTAT5 only increased in Treg cells, indicating CD4^+^ T cell subset-specific activation (Supplementary Fig. 1B). Following treatment, placebo recipients showed further increases in pSTAT1, pSTAT3, and pSTAT5 at T+1.5, consistent with responses to dying parasites (Fig. 1D, Supplementary Fig. 1C). In contrast, ruxolitinib caused a marked, but transient reduction in pSTATs at T+1.5 across all CD4^+^ T cell subsets (Fig. 1D, Supplementary Fig. 1D). By T+5, phosphoprotein levels returned to baseline in the placebo group but remained elevated in the ruxolitinib group, suggesting prolonged CD4^+^ T cell activation (Fig. 1D).

Cytokine stimulation assays yielded similar effects, whereby ruxolitinib significantly reduced IFNβ-induced pSTAT1, pSTAT3, and pSTAT5, as well as IL-2-induced pSTAT5, at T+1.5 (Fig. 1E, Supplementary Fig. 2A–D). Analysis of fold changes (T to T+1.5) revealed consistent clustering by treatment group, where placebo participants showed increased pSTAT expression, while ruxolitinib-treated participants showed reduced responses (Fig. 1F–G). One participant who received only two ruxolitinib doses (participant D722, treatment stopped due to as protocol-defined toxicity rules (Webster et al., 2025)) grouped with placebo rather than ruxolitinib-treated participants. While pSTAT1 and pSTAT3 responses were consistent across CD4^+^ T cell subsets, pSTAT5 showed more variability amongst CD4^+^ T cell subsets in unstimulated conditions (Fig. 1F). Similar patterns of phosphorylation were observed following IL-2 stimulation, although the effect was relatively uniform across all CD4^+^ T cell subsets (Supplementary Fig. 2C-E).

Beyond JAK/STAT activation, *P. falciparum* infection and treatment also affected other CD4^+^ T cell signalling pathways and pCREB, pERK, and pS6 levels declined following treatment in unstimulated and in PMA/ionomycin stimulated responses (Supplementary Fig. 1E, Supplementary Fig. 3A). However, for pCREB this reduction was significantly greater in placebo compared to ruxolitinib recipients (Supplementary Fig. 1F, Supplementary Fig. 3A). Although more variable than changes in pSTATs (Supplementary Fig. 3B), these changes indicate broader effects of *P. falciparum* infection on CD4^+^ T cell signalling pathways, and also suggest preservation of these pathways by ruxolitinib. Together, these findings demonstrate that ruxolitinib effectively suppresses JAK/STAT signalling in CD4^+^ T cells during malaria, dampening both *in vivo* activation and IFNβ/IL-2-induced responses, while it may preserve other mitogen-driven CD4^+^ T cell signalling pathways, albeit in a more heterogeneous way.

### Th1, Tfh and Tr1 cell responses are more rapidly activated in second infection and modulated by ruxolinitib treatment

To assess the impact of ruxolitinib on CD4^+^ T cell subset development and expansion, we performed high-dimensional surface phenotyping CyTOF analysis of PBMCs at multiple time points during the first and second infection (I, T, T+3, T+7, T+20; re-inoculation (rI), rT, rT+3, rT+7, end of study (EOS)) (Supplementary Fig. 4A). CD4^+^ T cells were identified within PBMCs (Supplementary Fig. 4B-C), and then clustered based on chemokine receptors and memory markers (CCR7, CXCR5, CXCR3, CCR6, CD45RA), regulatory markers (CD127, CD25, LAG3, CD49b), and activation markers (CD38, PD1, ICOS, HLA-DR), identifying 18 subsets (Fig. 2A-B).

**Figure 2:**
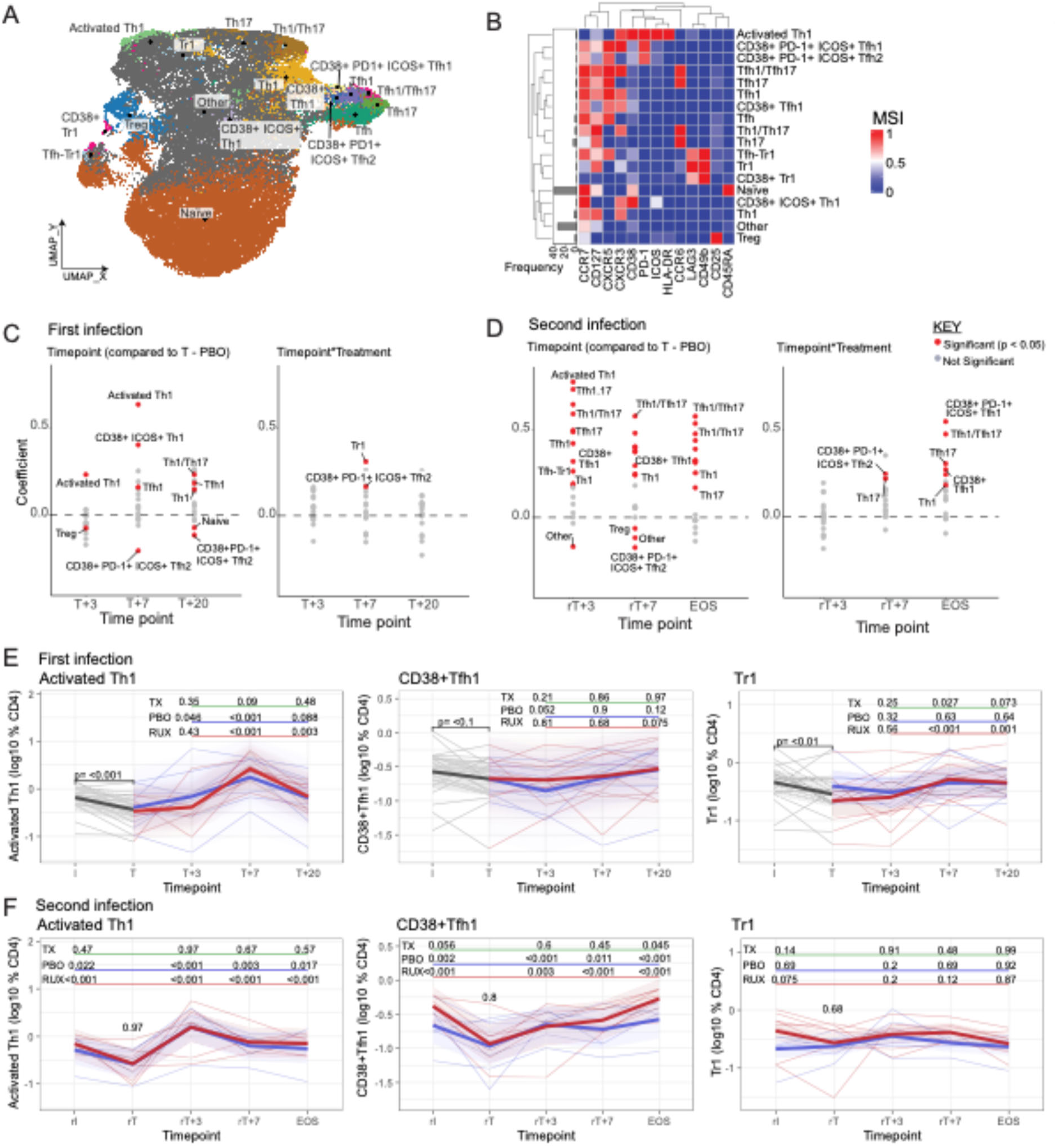
CD4 T cell subsets and activation in first and second infection. CD4 T cells were analysed by CyTOF at inoculation (I), treatment (T) and post treatment timepoints (T+3, T+7 and T+20) in first infection, and at re-inoculation (rI), re-treatment (rT) and post treatment timepoints (rT+3, rT+7, EOS) in second infection. **A)** UMAP and **B)** cluster marker expression of identified CD4 T cell subsets. **C/D)** Coefficient from mixed effect models analysing CD4 T cell subsets, analysing changes with time compared to treatment (T in first infection and rT in second infection), and the interaction between time and ruxolitnib treatment (Timepoint*Treatment). Significant changes are indicated in red. Coefficient of zero, indicating no change is marked by a dashed line. **E/F)** Activated Th1, CD38+ Tfh1 and Tr1 cells in first and second infection. Data are log10 frequencies of CD4 T cells, with thin lines representing each participant. All datapoints prior to randomisation are in grey; following randomisation ruxolitinib-treated participants are in red and placebo-treated are in blue. P values are from linear mixed effect models, with bold lines representing the mean of the predicted values from the fitted models for each group. TX is p values for the interaction term between each timepoint (compared to T) and treatment groups (underlined with green). P values for the comparison between each timepoint and T are shown for the placebo (PBO, underlined with blue) and ruxolitinib (RUX, underlined with red) groups, and were determined from contrasts. P value for the difference at rT between placebo and ruxolitinib group, calculated from the intercept is shown at rT. See also Supplementary Figure 4.

Longitudinal analysis revealed distinct CD4^+^ T cell dynamics in first and second infection, which were modulated by ruxolitinib (Fig. 2C-D). During the first infection, there was a reduced frequency of several CD4^+^ T cell subsets, including activated Th1 and Tr1 cells between inoculation and treatment (Fig. 2E-F, Supplementary Fig. 4D). This is consistent with other human studies that show transient reductions of circulating CD4⁺ T cell subsets during acute malaria, with recovery after treatment and accompanying evidence that lymphocytes relocate to secondary lymphoid tissues (Boyle et al., 2015b; Mandala et al., 2022; van Wolfswinkel et al., 2017). After treatment, activated Th1 cell frequency increased at both T+3 and T+7, consistent with strong CD4^+^ T cell activation (Fig. 2C). Activated Tfh2 cells decreased, and Tr1 cell frequencies remained low. Ruxolitinib modestly affected the frequencies of Tr1 and activated Tfh2 cells, but overall changes were small (Fig. 2C). In the second infection, multiple CD4^+^ T cell subsets, including activated Th1 and Tfh cell populations, expanded more rapidly, with the greatest changes at rT+3, consistent with a recall response. Unlike in the first infection, ruxolitinib significantly modulated these recall responses, including increased frequencies of Th1 cells and activated Tfh1 and Tfh2 cells (Fig. 2D). CD4^+^ T cell subset-specific kinetics were also evident (Fig. 2E-F, Supplementary Fig. 4D-E). For example, activated Th1 cells peaked at T+7 in the first infection but at rT+3 in the second, and were not affected by ruxolitinib. Similar trends were observed for CD38^+^ICOS^+^ Th1 cells (Supplementary Fig. 4D-E). CD38^+^ Tfh1 cells showed minimal change in the first infection but expanded at all post-treatment time points in the second infection, with markedly higher frequencies in ruxolitinib-treated participants at EOS. Similar kinetics were observed for CD38^+^ PD1^+^ ICOS^+^ Tfh1 and Tfh2 cells, which both also remained significantly elevated in ruxolitinib recipients in second infection at EOS (Supplementary Fig. 4D-E). Tr1 cells were detected at low frequency, possibly due to staining limitations of LAG3 and CD49b in CyTOF, with a modest ruxolitinib-dependent increase at T+7 during the first infection only (Fig. 2E-F). Together, these data indicate comparable memory Th1 cell responses across treatment groups, but a more robust and sustained germinal centre–associated response in ruxolitinib-treated participants during second infection, reflected in sustained Tfh cell subset frequencies.

### Ruxolitinib transiently delays induction of co-inhibitory rich Tr1 cell subsets during first infection

We and others have previously reported on the phenotypic heterogeneity of Tr1 cells and the impact this has on regulatory function (Brockmann et al., 2018; Edwards et al., 2023). We developed an optimised spectral flow cytometry panel to identify Tr1 cells and to characterise their phenotype in detail (Supplementary Fig. 5). To assess whether ruxolitinib influenced Tr1 cell phenotype and functional potential, we examined co-inhibitory receptors (CIR: CTLA4, CCR2, TIM3, CCR5, TIGIT, PD1), memory markers (CD45RA, CCR7, CXCR5, CXCR3, CD161), and activation/proliferation markers (Ki67, CD38, ICOS) using unbiased clustering (Fig. 3A-B). Ten Tr1 clusters were identified. Four clusters (C4, C5, C6, C10) were CIR-rich, expressing high levels of 4–5 CIRs, and overlapped with Tfh (CXCR5^+^) and Th1 (CXCR3^+^) cell phenotypes, consistent with PBMC data (Fig. 2). Cluster C4 also had elevated expression of Ki67, ICOS, and CD38 expression, indicating an activated and proliferating status (Fig. 3B).

**Figure 3:**
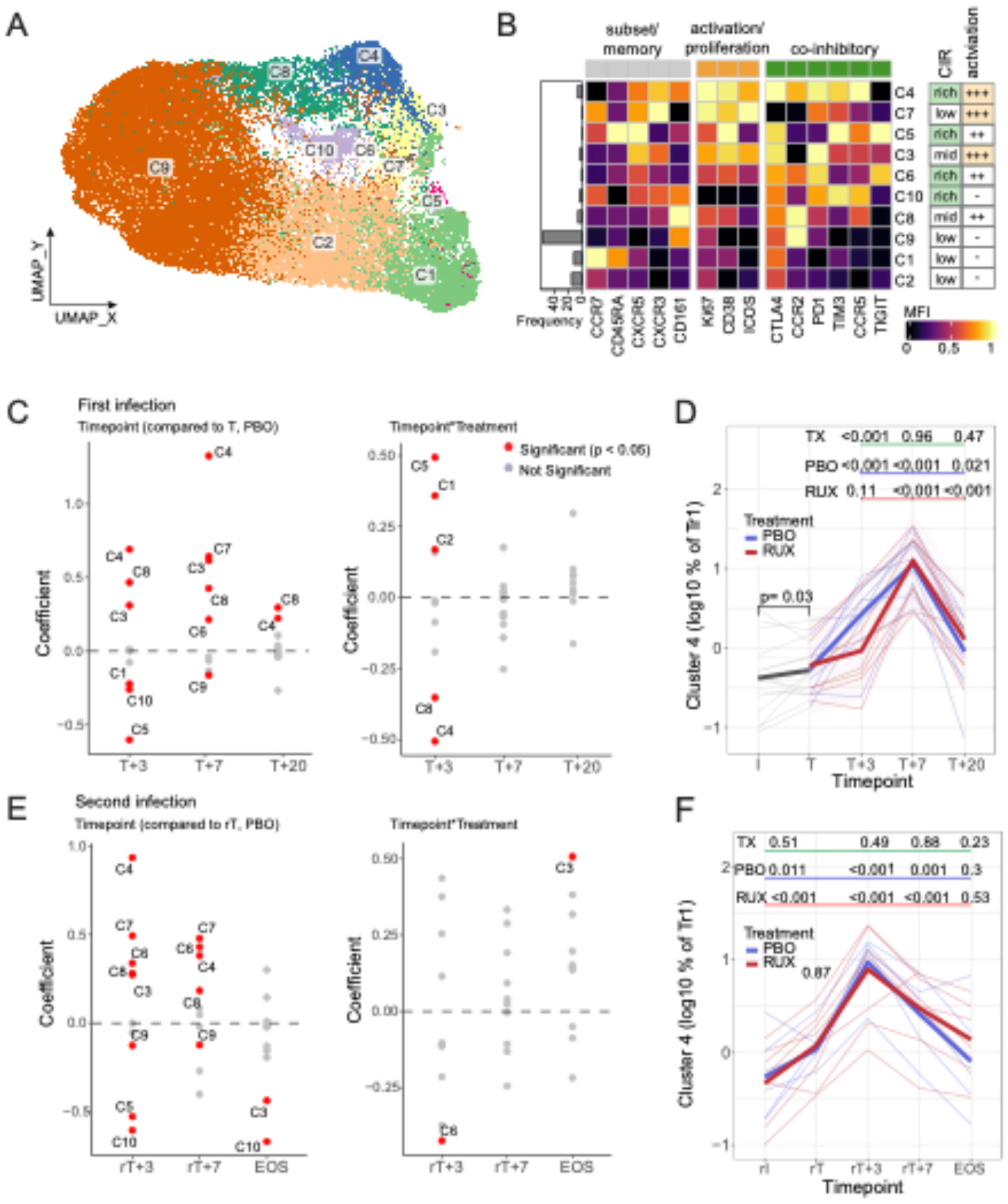
Induction of Tr1 co-inhibitory receptor rich CD4 T cells and impact of ruxolitinib. A/B) UMAP of Tr1 CD4 T cell clusters based on expression of subset, memory, activation, proliferation and co-inhibitory receptor (CIR) markers showed in heatmap. **C)** Expression of markers on subsets identified within Tr1 cells. CIR rich subsets are indicated. Subsets identified were analysed in mixed models in first and second infection as a proportion of total Tr1 cells. **D/F)** Coefficient from mixed effect models analysing Tr1 cell subsets, analysing changes with time compared to treatment (T in first infection and rT in second infection), and the interaction between time and ruxolitnib treatment (Timepoint*Treatment). Significant changes are indicated in red. Coefficient of zero, indicating no change, is marked by a dashed line. **A/E/G)** Data are log10 frequencies of CD4 T cells, with thin lines representing each participant. Prior to randomisation data are in grey; following randomisation ruxolitinib-treated participants are red and placebo-treated participants are blue. P values are from linear mixed effect models, with bold lines representing the mean of the predicted values from the fitted models for each group. TX is p values for the interaction term between each timepoint (compared to T) and treatment groups. p values for the comparison between each timepoint and T are shown for the placebo (PBO) and ruxolitinib (RUX) groups, which were determined from contrasts. P value for the difference at rT between placebo and ruxolitinib group, calculated from the intercept is shown at rT. See also Supplementary Figure 5.

During first infection, multiple Tr1 clusters expanded, including CIR-rich and activated C4 and C6 clusters at T+3 and T+7, alongside expansion of an activated cluster (C8). In contrast, naïve-like clusters (C1, C5; CD45RA^+^) or non-activated clusters (C10) contracted.

Importantly, the expansion of the most CIR-rich and activated cluster, C4, was significantly reduced in ruxolitinib-treated participants at T+3, and this effect was transient, with frequencies equalising by T+7 (Fig. 3C-D). In contrast, naïve, and non-activated clusters (C1, C2 and C5) were relatively expanded in ruxolitinib-treated participants at T+3 (Fig. 3C). In second infection, the highly activated CIR-rich C4 cluster (C4) re-expanded, peaking at rT+3, and expansion was not influenced by ruxolitinib treatment in first infection (Fig. 3E-F). Together, these results show that ruxolitinib transiently suppresses the induction of Tr1 cell subsets with high activation and inhibitory potential during first infection but does not show a long-term effect on the capacity of Tr1 cells to expand during a second infection.

### Effects of ruxolitinib on antigen-specific CD4^+^ T cell responses during first and second infection

To examine malaria-specific CD4^+^ T cell responses, we performed Activation-Induced Marker (AIM) assays with pRBC stimulation on samples collected during first and second infection (I, T, T+3, T+7, T+20; rI, rT, rT+3, rT+7, EOS). Malaria-specific CD4^+^ T cells were identified by OX40 and CD69 expression (AIM^+^ cells), and subsets defined using CXCR5, PD1, CXCR3, CCR6, LAG3, CD49b, and CCR4 (Fig. 4A, Supplementary Fig. 6). Identified subsets included Tr1 (LAG3^+^ CD49b^+^), Tfh (CXCR5^+^ PD1^+^, LAG3^-^ CD49b^-^), Th1 (CXCR3^+^ CCR4^−^ CCR6^−^), Th2 (CXCR3^−^ CCR4^+^ CCR6^−^), and Th17 (CXCR3^−^ CCR4^−^ CCR6^+^) cells. Within Tr1 cells, Tfh.Tr1 cells (CXCR5^+^ PD1^+^) were also analysed, and Tfh cell subsets were further classified (Tfh1, Tfh2, Tfh17), as for Th1, Th2 and Th17 cell lineages.

**Figure 4:**
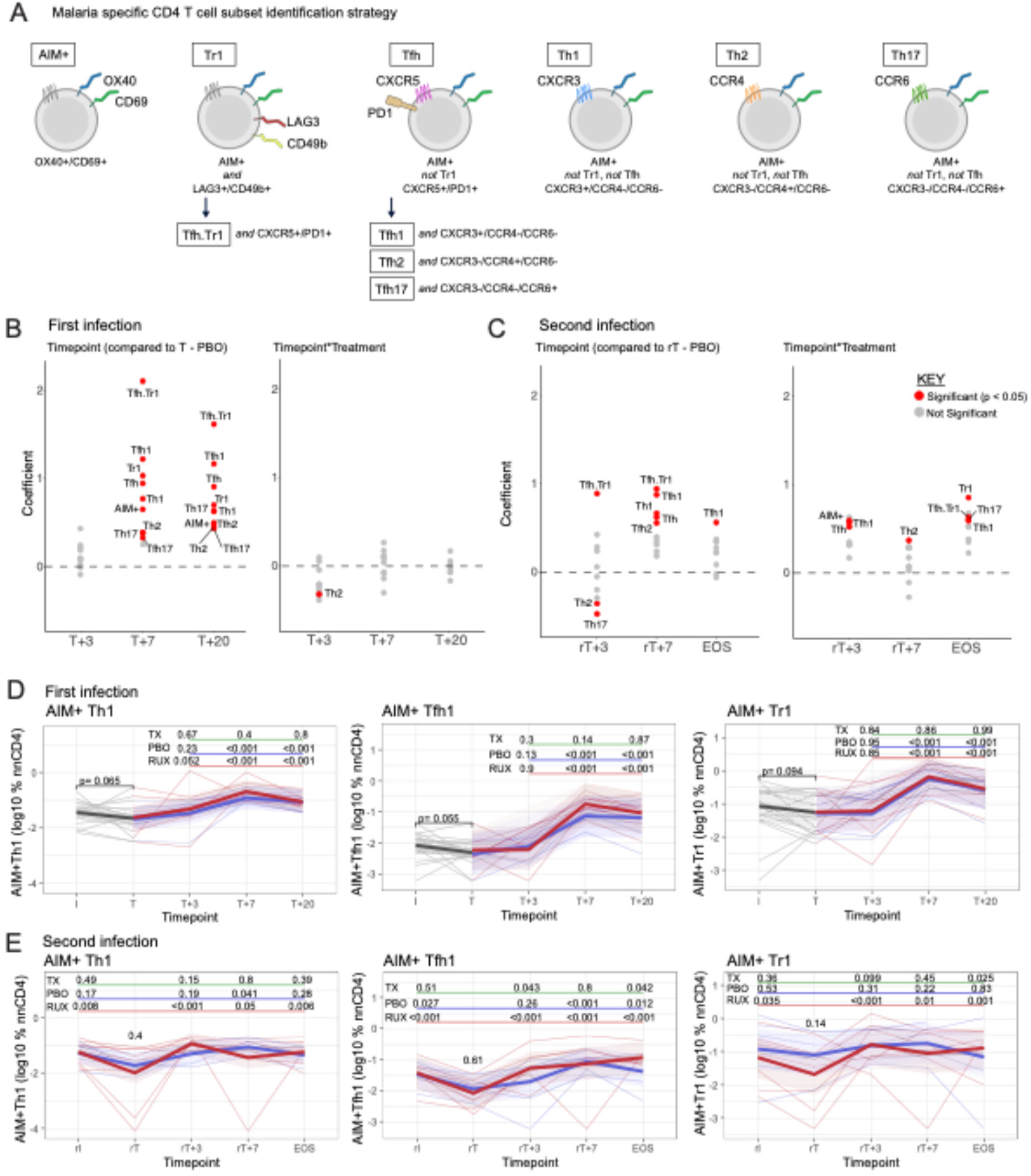
Malaria specific CD4 T cell responses during first and second infection. Parasite specific CD4 T cells were identified using Activation Induced Marker assay at inoculation (I), treatment (T) and post treatment timepoints (T+3, T+7 and T+20) in first infection, and at re-inoculation (rI), re-treatment (rT) and post treatment timepoints (rT+3, rT+7, EOS) in second infection. **A)** Schematic of identified CD4 T cell subsets. **B/C)** Coefficient from mixed effect models analysing CD4 T cell subsets, analysing changes with time compared to treatment (T in first infection and rT in second infection), and the interaction between time and ruxolitnib treatment (Timepoint*Treatment). Significant changes are indicated in red. Coefficient of zero, indicating no change is marked by a dashed line. **D/E)** AIM+ Th1, AIM+ Tfh1 and AIM+ Tr1 cells in first and second infection. Data are log10 frequencies of malaria specific AIM+ CD4 T cells, with thin lines representing each participant. Prior to randomisation is in grey, following randomisation ruxolitinib-treated participants are red and placebo-treated are blue. P values are from linear mixed effect models, with bold lines representing the mean of the predicted values from the fitted models for each group. TX is p values for the interaction term between each timepoint (compared to T) and treatment groups (underlined in green). P values for the comparison between each timepoint and T are shown for the placebo (PBO, underlined in blue) and ruxolitinib (RUX, underlined in red) groups, which were determined from contrasts. P value for the difference at rT between placebo and ruxolitinib group, calculated from the intercept is shown at rT. See also Supplementary Figure 6 & 7.

In the first infection, AIM^+^ CD4^+^ T cells increased significantly at T+7 and T+20 across all subsets (Fig. 4B). Induction of AIM^+^ CD4^+^ T cells was comparable between placebo- and ruxolitinib-treated participants, consistent with findings in the total CD4^+^ T cell compartment (Fig. 2). By the time of second infection (rI), AIM^+^ CD4^+^ T cell frequencies had returned to baseline, except for AIM^+^ Tfh1 cells, which remained elevated (Supplementary Fig. 7A).

Following the second infection, CD4^+^ T cell AIM^+^ responses re-emerged but were restricted to Th1, Tfh, and Tr1 (notably Tfh.Tr1) cell subsets, indicating a more focused recall response (Fig. 4C). In contrast to the first infection, ruxolitinib-treated participants showed a significantly stronger and faster recall response, particularly within the Tfh cell compartment, and these responses remained elevated compared to placebo at EOS (Fig. 4C).

Analysis of CD4^+^ T cell subset frequencies over time revealed that in the first infection, AIM^+^ Th1, Tfh, and Tr1 cell frequencies peaked at T+7 and remained elevated at T+20 (Fig. 4D), while AIM^+^ Th2 and Tfh2 peaked later at T+20 (Supplementary Fig. 7B). During the second infection, an earlier and more sustained antigen-specific responses were observed in ruxolitinib-treated participants. The frequencies of total AIM^+^ CD4^+^ T cells, as well as AIM^+^ Th1, Tfh1, Tr1 and total Tfh cells were significantly higher in the ruxolitinib group at rT+3 and EOS, compared with placebo (Fig. 4C & E, Supplementary Fig. 7C). Together, analyses of both the total and antigen-specific CD4^+^ T cell compartments indicate that while ruxolitinib had limited observable effect on responses during the first malaria infection, its administration at first infection enhanced the magnitude and duration of the CD4^+^ T cell recall response following a second infection, most notably within the Tfh cell lineage.

### Th1, Tfh and Tr1 cells share overlapping transcriptional profiles and emerge rapidly after anti-parasitic drug treatment following second infection

To better understand the impact of ruxolitinib on primary and recall CD4^+^ T cell responses, we used scRNA/TCRseq to transcriptionally profile responses in 8 participants during first and second infection (n=4 placebo, n=4 ruxolitinib, 10 timepoints each) (Fig. 5A). Antigen experienced CD4^+^ T cells (CD45RA^-^) were sorted by flow cytometry and analysed, and malaria-specific cells were also identified by AIM assays, sorting CD69^+^/OX40^+^ cells for sequencing (Fig. 5A, Supplementary Fig. 8A-B). scRNAseq data from antigen-experienced CD4^+^ T cells and AIM^+^ CD4^+^ T cells were integrated based on donor (Fig. 5B). The unstimulated antigen-experienced CD4^+^ T cells were then selected for downstream clustering and annotation. Ten CD4^+^ T cell clusters were identified from 127,482 high quality unstimulated cells (Fig. 5C, Supplementary Fig. 8C-E).

**Figure 5:**
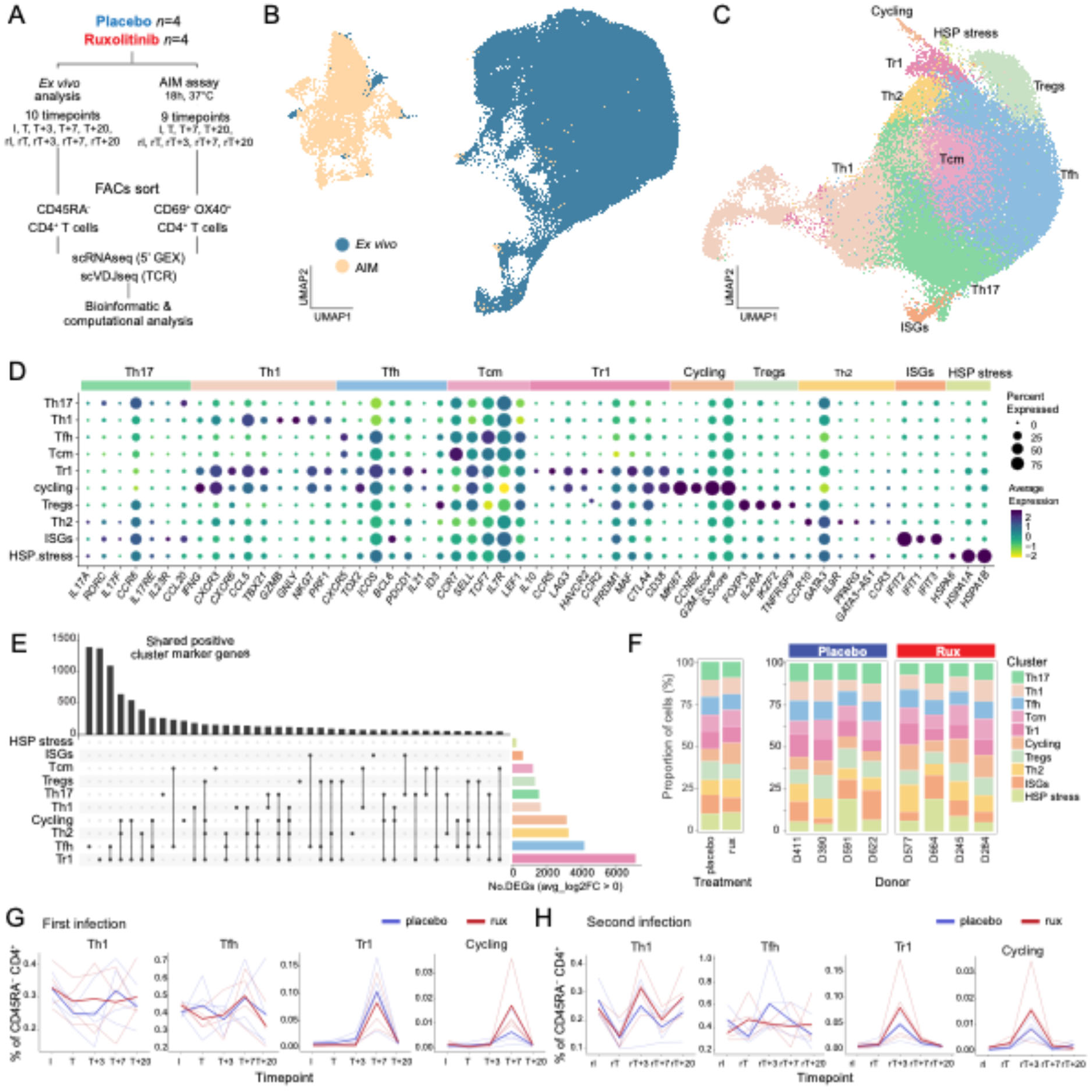
scRNAseq reveals overlapping Th1, Tfh and Tr1 transcriptional profiles. **A)** Experimental design showing scRNA/TCRseq profiling of antigen experienced (CD45RA^-^) CD4^+^ T cells (Ex vivo) and malaria specific (CD69^+^OX40^+^) CD4^+^ T cells identified by AIM assays (AIM) from 8 participants (n = 4 placebo, n = 4 ruxolitinib) across 9 – 10 timepoints during first and second infection. **B)** UMAP visualisation showing the Ex vivo and AIM cells after integration by donor. **C)** UMAP visualisation showing Ex vivo cells after subsetting and performing unsupervised clustering analysis. **D)** Dot plot showing the average expression of curated marker genes for each Ex vivo cluster. **E)** Upset plot showing the number of shared marker genes between Ex vivo clusters. The bar graph on the right shows the total number of differentially expressed cluster marker genes for each subset. **F)** Stacked bar plots showing the proportion of cells within each cluster by treatment and donor. **G)** Line graphs showing the cluster frequency as a percentage of total Ex vivo CD4^+^ T cells during the first and **H)** second infections. Lines are coloured by treatment group, with bold lines representing group means and transparent lines showing individual donor values. See also Supplementary Figures 8 & 9.

CD4^+^ T cell clusters included Th1, Tfh and Tr1 cell subsets. Th1 cells were identified based on elevated *CXCR3, CCL5, TBX21* (encoding T-bet), and cytotoxic markers *NKG7, GNLY, GZMB* and *PRF1;* Tfh cells could be identified based on elevated *CXCR5*, *TOX2*, *ICOS*, and *BCL6*; and Tr1 cells could be identified based on elevated *IL10*, *CCR5*, *LAG3*, *HAVCR2* (encoding Tim3), and *PRDM1* (encoding BLIMP1) (Fig. 5D). Consistent with the overlapping phenotypes of Tr1 cells with Th1 and Tfh cells (Fig. 2, 3 & 4), the Tr1 cell cluster also expressed high levels of Th1 cell markers including *IFNG, CXCR3, TBX21* and Tfh cell markers including *CXCR5* and *BCL6* (Fig. 5D). A cycling cell cluster was also identified based on expression of *MKI67 (*encoding Ki67), which also expressed high levels of *CD38* consistent with activation. This cycling cluster shared gene expression profiles with Th1 (including *IFNG* and *CXCR3)*, Tfh (including *TOX2* and *IL21*) and Tr1 (including *LAG3* and *HAVCR2)* cells (Fig. 5D). The shared gene expression by Th1, Tfh and Tr1 cells was also confirmed by analysis of all cluster marker genes, which identified 1098 overlapping marker genes between Tfh and Tr1 cells, 186 genes between Th1 and Tr1 cells and 270 genes between cycling, Tfh and Tr1 cells (Fig. 5E).

In addition to the Th1, Tfh, and Tr1 cell clusters, several other CD4⁺ T cell subsets were identified. Th17 cells were defined by expression of *CCR6* and *RORC*; central memory T (Tcm) cells by expression of *CCR7*; regulatory T (Treg) cells by expression of *FOXP3* and *IL2RA*; and Th2 cells by expression of *GATA3*. Two smaller clusters were also detected: one characterised by high expression of type I interferon–stimulated genes (ISGs), including *IFIT2*, *IFIT1*, and *IFIT3*; and another marked by elevated heat shock protein expression (HSP stress cluster) (Fig. 5C-D). The distribution of these CD4^+^ T cell clusters varied by donor, treatment group and timepoint (Supplementary Fig. 8F-H).

Further sub-clustering of Th1, Tfh and Tr1 cells revealed substantial heterogeneity within each subset. Tr1 cells segregated into three distinct transcriptional clusters: a co-inhibitory receptor–rich cluster characterised by elevated *CCR2*, *CCR5*, *LAG3*, and *HAVCR2* (encoding TIM-3) expression; a memory-like cluster defined by *CXCR5* and *CCR7* expression; and a proliferating cluster marked by *MKI67* expression (Supplementary Fig. 9A-B). The proportions of these Tr1 cell sub-clusters varied across timepoints, with the CIR-rich cluster relatively reduced at T+7 in first infection in ruxolitinib-treated participants and with all three appearing more frequent in ruxolitinib-treated participants during the second infection at the rT+3 timepoint (Supplementary Fig. 9C). Tfh cells were further divided into six clusters: a Tfh/Th17 cell-like cluster, a Tfh cell effector cluster, a Tfh cell activated cluster, a Tfh cell AREG⁺ cluster, a Tfh cell stress cluster, and a Tfh cell ID3⁺ cluster (Supplementary Fig. 9D-E). The frequencies of these clusters also varied with timepoint and treatment. Notably, the Tfh activated cell cluster, with elevated expression of *CD38* and *HLA-DRA*, was increased in ruxolitinib-treated participants following second infection at the late timepoint of rT+20, consistent with enhanced activation or persistence of Tfh cell responses (Supplementary Fig. 9F). Th1 cells could be divided into six clusters, including three cytotoxic clusters distinguished by expression of *GNLY*, *GZMB*, *CCL4*, and *CCL3*; two Th1/Th17 cell-like clusters expressing *CCR6* and *CXCR3*; and a Th1/Tfh/Tr1 cell-like cluster expressing *IL10*, *CXCR5* and *CXCR3* (Supplementary Fig. 9G-H). The distribution of Th1 cell sub-clusters also differed across timepoints and between treatment groups (Supplementary Fig. 9I).

While frequencies of CD4^+^ T cell clusters differed across participants (Fig. 5F), there was a clear expansion of Th1, Tfh and Tr1 cells in response to malaria. Following first infection, Tr1, Tfh and cycling cells expanded at T+7; these appeared higher in ruxolitinib-treated participants for cycling cells (Fig. 5G; Supplementary Fig. 9I). In second infection, the re-emergence of cycling, and Tr1, Tfh, Th1 cell subsets occurred earlier in infection at rT+3, consistent with CyTOF and flow cytometry data (Fig. 2 & 3). A robust recall response was also measured during second infection, particularly in cycling cells from ruxolitinib-treated participants (Fig. 5H, Supplementary Fig. 9J). Thus, these results support earlier findings from CyTOF and flow cytometry analysis and reinforce the impact of ruxolitinib treatment during first infection on modulating the expansion and balance of Th1, Tr1 and Tfh cells during second infection. Furthermore, transcriptional overlap between Th1, Tr1 and Tfh cells was observed, suggesting plasticity in these CD4^+^ T cell subsets.

### Ruxolitinib treatment promotes a pro-inflammatory gene signature in CD4^+^ T cells

We next examined the hallmark IFN-alpha response gene signature across CD4⁺ T cell subsets between ruxolitinib- and placebo-treated participants. Across all subsets, ruxolitinib treatment was associated with significantly reduced IFN-alpha response scores at T+3, consistent with suppression of type I IFN–driven pathways by ruxolitinib treatment (Fig. 6A). To determine the impact of ruxolitinib on the transcriptional landscape of Th1, Tfh, and Tr1 cells, we performed pseudobulk differential gene expression analysis between ruxolitinib-and placebo-treated cells (Fig. 6B). This analysis was performed at T+7 during first infection and rT+3 following the second infection, as these timepoints represented peak CD4^+^ T cells responses (Fig. 2-5). We identified 275 differentially expressed genes (DEGs) at T+7 (147 upregulated in ruxolitinib, 128 in placebo) and 262 DEGs at rT+3 (143 up in ruxolitinib, 119 in placebo) in Tr1 cells. Genes upregulated in ruxolitinib-treated Tr1 cells included cytotoxic genes such as *GZMA*, *GZMH*, *NKG7*, and *GNLY*, suggesting relatively enhanced cytotoxic potential (Fig. 6B). Pathway analysis indicated enrichment of the Interferon Gamma Response pathways in ruxolitinib-treated participants during first and second infection, along with other inflammatory pathways (Fig. 6C). Tfh cell analysis revealed 434 DEGs at T+7 (234 up in ruxolitinib, 200 in placebo) and 338 DEGs at rT+3 (194 up in ruxolitinib, 144 in placebo) (Fig 6B). Again, pathway analysis highlighted Hallmark Interferon Gamma Response pathways enrichment in Tfh cells from ruxolitinib-treated donors during first and second infection (Fig 6C). We also identified 182 DEGs at T+7 (118 up in ruxolitinib, 64 in placebo) and 178 DEGs at rT+3 (105 up in ruxolitinib, 73 in placebo) in Th1 cells (Fig. 6B), and as expected for this CD4^+^ T cell subset, Hallmark Interferon Gamma Response pathways were enriched in ruxolitinib Th1 cells at both timepoints (Fig. 6C). Overall, these results indicate that ruxolitinib drives antigen-specific CD4^+^ T cell subsets towards a more pro-inflammatory transcriptional signature that is maintained following re-exposure to parasite antigen.

**Figure 6:**
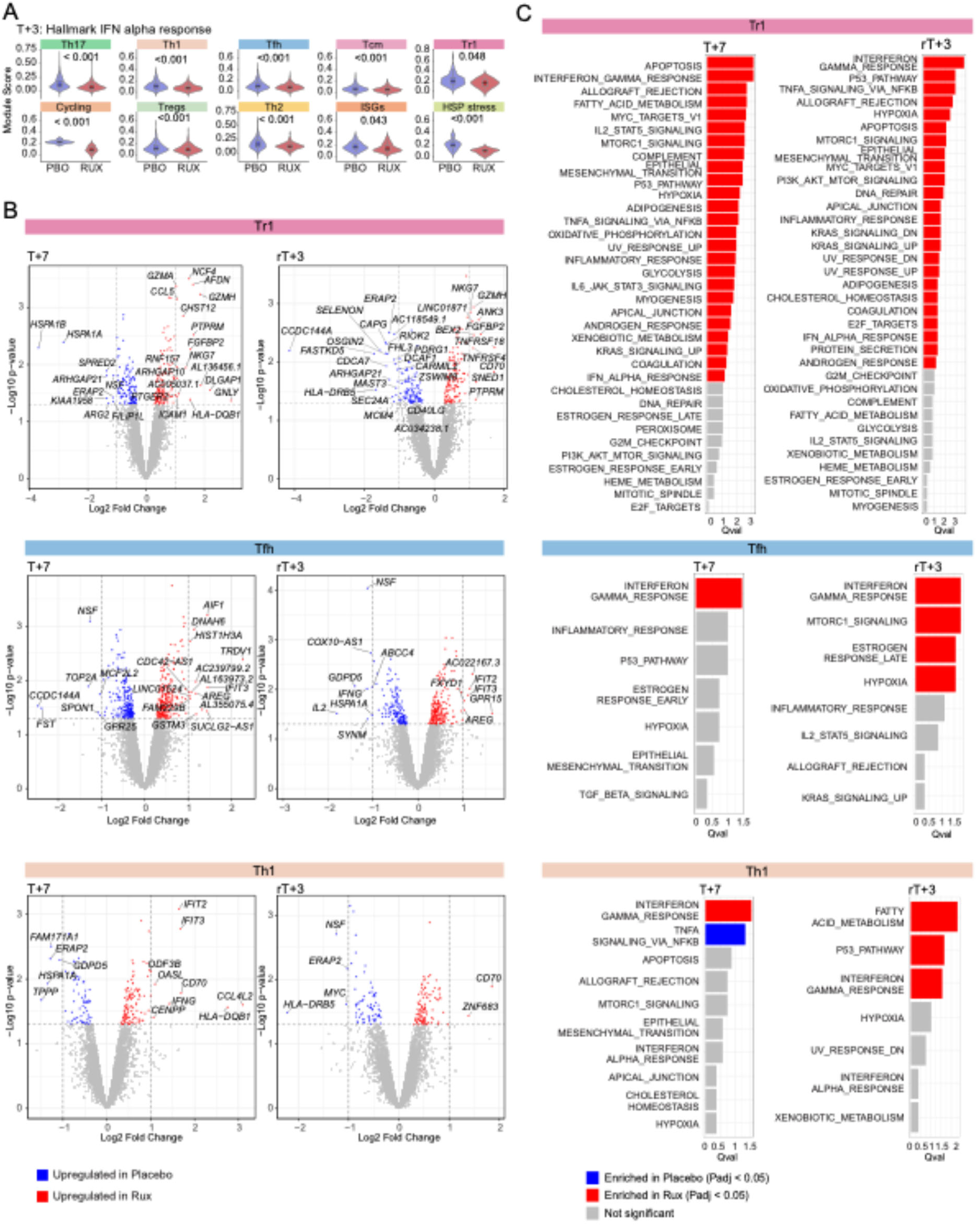
Ruxolitinib treatment promotes transcriptional programs associated with enhanced CD4^+^ T cell function. **A)** Violin plots showing the Hallmark IFN alpha response module score for each CD4^+^ T cell subset at T+3 by treatment group. Boxplots are overlaid on the violins, with the middle line representing the median and the box edges indicating the 25^th^ and 75^th^ percentiles. Statistical comparison between treatment groups was performed using a Wilcoxon rank-sum test. **B)** Volcano plots showing pseudobulk edgeR differential gene expression analysis comparing ruxolitinib and placebo treatment groups at T+7 and rT+3 within the Tr1, Tfh and Th1 cell clusters. Genes with P<0.05 are highlighted, blue indicates upregulation in placebo-treated and red indicating upregulation in ruxolitinib-treated groups. Genes with log2 fold change >1 are labelled. **C)** Waterfall plots showing single cell pathway analysis (SCPA) for Tr1, Tfh and Th1 cells at T+7 and rT+3 using the Hallmark gene sets.

### Th1, Tfh and Tr1 cells share clonal relationships and can be tracked over the course of first and second infection

Finally, to dissect the relationships and expansion of the Th1, Tfh and Tr1 cells at the clonal level within *P. falciparum*-specific cells, we examined CD4^+^ T cell subsets and clonal dynamics within the AIM assay dataset. AIM^+^ cells were selected from the integrated scRNAseq dataset (Figure 5B) and taken forward for clustering and cell annotation. Clustering of AIM^+^ cells identified eight transcriptionally distinct CD4^+^ T cell subsets; Th1 cells were defined based on expression of *IFNG*, *GNLY*, *GZMK*, *GZMB*; Tfh cells by expression of *CXCR5*, *ICOS* and *CXCL13*; Tr1 cells by expression of *IL10*, *LAG3*, and *PDCD1*. As seen in unstimulated CD4^+^ T cells, there was significant sharing of transcriptional signatures across Th1, Tfh and Tr1 cells, with Tr1 cells co-expressing Th1-and Tfh cell-associated transcripts, including *IFNG*, *GZMB*, and *ICOS*, *CXCL13* (Fig. 7A-B, Supplementary Fig. 10A-C). Other identified CD4^+^ T cell subsets included: Th17 cells defined by expression of *IL22* and *IL17A*; T regulatory (Treg) cells that expressed *FOXP3*; Th2 cells identified by expression of *IL4* and *IL5;* a cluster enriched for type I IFN stimulated genes (annotated ISGs) including *CXCL10*, *IFI27* and *IFIT1;* and a cluster expressing *KLF2*, *TLR3*, *ALOX5*, consistent with an activated CD4^+^ T cell phenotype (Fig. 7A-B). The relative expansion of Th1, Tfh and Tr1 cells in response to *P falciparum* infection was detected at T+7 and subsequent timepoints (Supplementary Fig. 10D).

**Figure 7:**
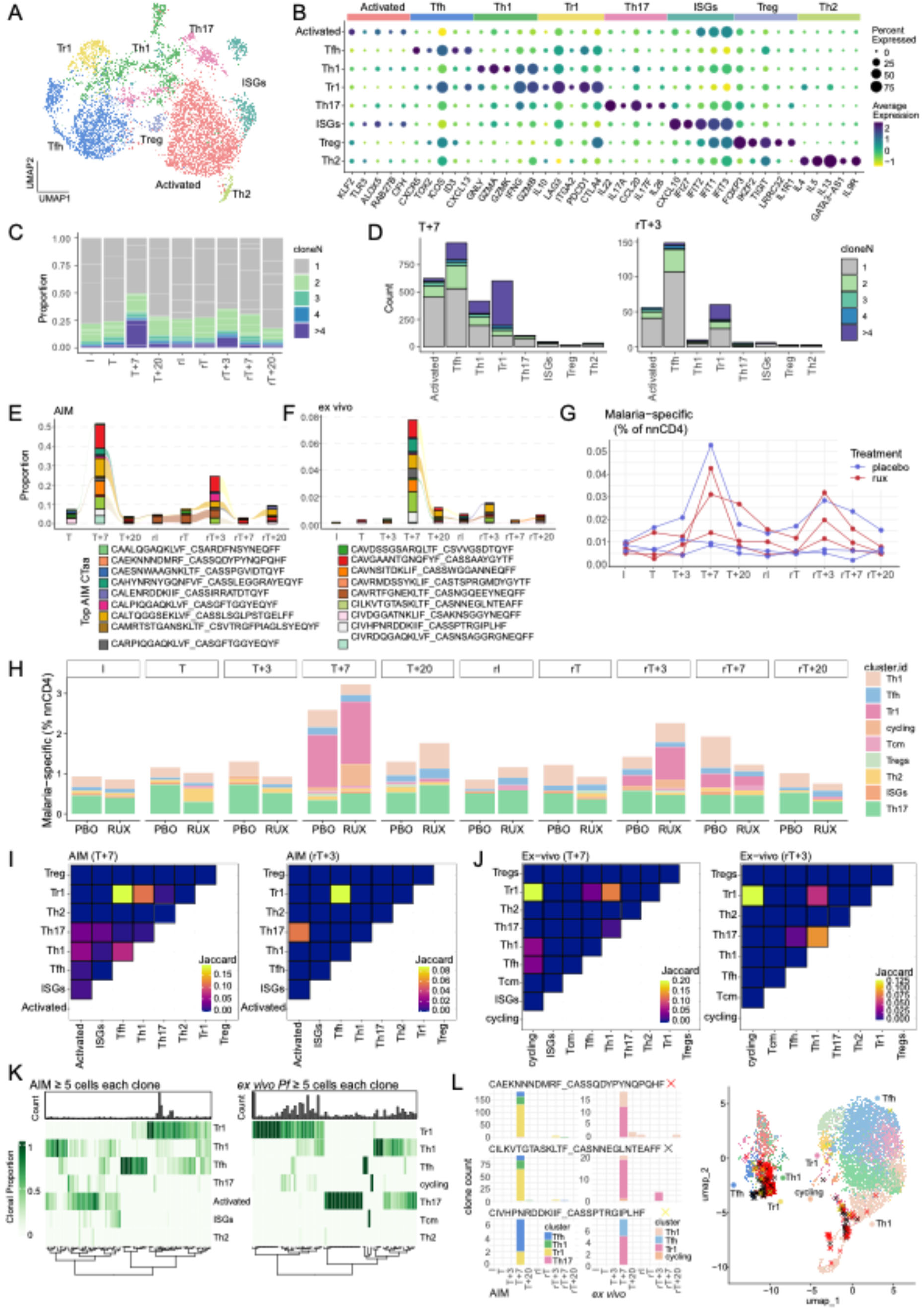
TCR clonal sharing between Th1-Tfh-Tr1 cells in primary and recall responses. **A)** UMAP visualisation showing the AIM cells after integration by donor. **B)** Dot plot showing the average expression of curated marker genes for each AIM cell cluster. **C)** Proportion of cells in each clone family size at each timepoint. **D)** Clonal family size count over time in each subset. **E/F)** Top 3 expanded P. falciparum-specific clones from each participant identified in AIM data set, in AIM **(E)** and within the ex vivo **(F)** data sets overtime. **G)** Frequency of P. falciparum-specific cells identified within ex vivo data set, for each participant by treatment group. **H)** P. falciparum-specific cells within ex vivo data set across time, within each subset by treatment group. **I/J)** Clonal overlap amongst P. falciparum-specific clones within AIM (**I)** or ex vivo **(J)** data sets expressed as Jaccard index. **K)** Clonal subset fidelity of highly expanded clones (≥5 cells per clone). Individual clones are in each column with distribution across subsets shown for AIM^+^ (left) and ex vivo (right) data. **L)** The three top expanded P. falciparum-specific clones are shown with subset fate indicated in bars across timepoints for AIM^+^ and ex vivo data, as indicated (left) or on the UMAP of integrated AIM^+^ and ex vivo data (right). See also Supplementary Figure 10.

Within the *ex vivo* and AIM^+^ data sets, complete TCRA/B pairs were captured for 62% of cells (88,276 cells, 73,514 unique clonal types). Within the AIM^+^ data set, clonal expansion was most evident at T+7 and rT+3, consistent with major activation of parasite-specific CD4^+^ T cells at these timepoints in first and second infection (Fig. 7C). At these two timepoints, clonal expansion was seen across CD4^+^ T cell subsets, but was highest in Tr1, Tfh and Th1 cells, consistent with these being dominant *P. falciparum*-specific CD4^+^ T cell subsets (Fig. 7D). Expanded clones in the AIM^+^ data set were detected across donors and could be traced between first and second infection (Fig. 7E). Using an antigen-specific, personalised TCR reference database approach (Marina-Zárate et al., 2025), expanded clones (≥2 cells with shared TCRA/B sequences) identified within the AIM^+^ dataset were classified as *P. falciparum*-specific. We then leveraged these clonotypes to mine the unstimulated *ex vivo* TCR dataset, allowing identification of malaria-specific clones without perturbing cell transcriptional state. This approach detected 1110 *P. falciparum-*specific CD4^+^ T cells in the *ex vivo* data, across 455 unique clones; these could be tracked across time and between first and second infections (Fig. 7F). Within *ex vivo* data, clonal expansion of these parasite-specific clones was largest at T+7 and rT+3 across donors (Fig. 7G-H). Further, these expanded clones were dominated by Th1, Tfh, Tr1 and cycling cell subsets particularly at T+7 and rT+3, with indications these *P. falciparum*-specific, clonally expanded cells were higher in ruxolitinib-treated participants (Fig. 7G, Supplementary Fig. 10E). Th17 cells accounted for a substantial proportion of AIM^+^ cells across all timepoints, regardless of treatment (Fig. 7H). However, given the existence of these cells prior to malaria infection and limited change in their frequency over the course of first and second infection, it is likely they represent cytokine responsive, rather than antigen-specific cells, as recently reported in AIM assays for SARS-CoV-2- and influenza-specific CD4^+^ T cells (Zheng et al., 2025). Within parasite-specific clones, Jaccard index analysis indicated there was clear evidence of clonal sharing across Th1, Tfh and Tr1 cells, and within *ex vivo* data, clones within these subsets overlapped with cycling cells (Fig. 7I-J). Indeed, analysis of clonal subset fidelity indicated that amongst the most highly expanded *P. falciparum-*specific clones (≥5 cells per clone), clones were detected across Tr1, Th1 and Tfh cell subsets in both AIM^+^ and *ex vivo* data sets (Fig. 7K). This finding was further supported by analysis of individual clones in both data sets, where individual clones were detected in multiple clusters in both AIM^+^ or *ex vivo* data sets, either at a single timepoint, or across time (Fig. 7L). Taken together, these data reveal enhanced expansion of malaria-specific CD4^+^ T cells following a second infection and plasticity within Th1, Tfh and Tr1 cells.

## Discussion

This study provides a comprehensive phenotypic and transcriptional assessment for how transient JAK1/2 inhibition with ruxolitinib reshapes human CD4⁺ T cell immunity during controlled human malaria infection. Data reveals that host-directed therapy can alter the quality, kinetics, and durability of anti-parasitic CD4⁺ T cell responses. Although ruxolitinib transiently suppressed type I IFN signalling and delayed the emergence of highly activated Tr1 cell subsets during first infection, this limited suppression did not impair the development of anti-parasitic immunity. Instead, ruxolitinib treatment altered the CD4⁺ T cell response, preserving broader signalling pathways, enhancing formation and recall of germinal centre-associated Tfh cells, and driving more pro-inflammatory transcriptional programs across Th1, Tfh and Tr1 cells. Clonal tracking demonstrated that these effects translated into more robust expansion and sustained persistence of *P. falciparum*-specific CD4⁺ T cell clones following a second infection. These findings show that short-course JAK/STAT inhibition can modulate inflammatory signals during acute infection while simultaneously augmenting the quality of immunological recall responses, thus identifying a promising strategy to stimulate better and more durable vaccine or drug-induced protection against malaria.

A primary goal of our host-directed strategy was to transiently limit the development of Tr1 cells in volunteers infected with *P. falciparum* following anti-parasitic drug treatment in a first infection. We and others have previously reported that co-inhibitory receptor expression among Tr1 cells is highly variable, both in terms of receptor type and expression level (Brockmann et al., 2018; Edwards et al., 2023). We found that the development of co-inhibitory receptor-rich Tr1 cells was most affected by ruxolitinib treatment. This was not unexpected given the existence of an autologous, type I IFN-dependent pathway for Tr1 cell development, which regulates co-inhibitory receptor expression and IL-10 production (Wang et al., 2023). The therapeutic effect of ruxolitinib on target type I IFN signalling in children with interferonopathies (Fremond et al., 2016) and in patients with dermatomyositis (Ladislau et al., 2018) supports our findings and extends the impact of this drug to type I IFN-mediated Tr1 cell development. The transient nature of this effect further suggests that transiently delaying Tr1 cell expansion does not increase the risk of excessive inflammation, supported by our clinical observations (Webster et al., 2025).

The spleen is a key secondary lymphoid tissue during malaria and is composed of the red pulp, which filters blood and removes damaged and infected erythrocytes, and the white pulp, which orchestrates adaptive immune responses (Mebius and Kraal, 2005). Within the white pulp, periarteriolar lymphoid sheaths (PALS) enriched for T cells, B cell follicles that initiate germinal centre reactions, and the marginal zone that captures blood-borne antigens, together form a specialised, compartmentalised architecture that supports coordinated immune activation (Bronte and Pittet, 2013). This organisation is maintained by an intricate network of stromal cells, including fibroblastic reticular cells, follicular dendritic cells, marginal zone stromal cells, and endothelial cells, which provide essential structural scaffolding, organise chemokine gradients such as CCL19, CCL21, and CXCL13, and guide the migration, positioning, and interactions of lymphocytes and antigen-presenting cells (Cupedo et al., 2004; Mueller and Germain, 2009). During acute inflammation, however, excessive cytokine production, tissue-remodelling enzymes, and altered chemokine expression can destabilise these stromal networks, collapse microanatomical niches, and disrupt the boundaries between T and B cell zones (Auffray et al., 2007). In malaria, parasites accumulate in the spleen (Kho et al., 2021), and inflammatory remodelling can impair cell trafficking, weaken germinal centre formation, and reduce the efficiency of T and B cell cooperation (Cadman et al., 2008; Ryg-Cornejo et al., 2016; Urban et al., 2005), underscoring the importance of preserving splenic architecture for effective anti-parasitic immunity. In this context, our findings suggest that in addition to its direct suppression of CD4⁺ T cell signalling and Tr1 cell differentiation, ruxolitinib may act indirectly by stabilising the lymphoid microenvironment. The capacity of ruxolitinib to reduce acute systemic inflammation during first infection (Webster et al., 2025), likely dampens inflammatory stress on splenic stromal cells, thereby preserving the structural and chemokine cues required for optimal Th1 and Tfh cell support, as has been observed in other inflammatory settings (Alexandre and Mueller, 2018; Gommerman and Browning, 2003). Acute malaria imposes substantial metabolic and transcriptional constraints on T cells within secondary lymphoid tissues, limiting their expansion, survival, and differentiation (Boyle et al., 2024; Vijay et al., 2020). By transiently reducing acute inflammatory responses, ruxolitinib may mitigate this disruption, maintaining functional splenic niches that enable enhanced cycling, prolonged activation, and increased clonal persistence of Th1 and Tfh cells during second infection. This model reconciles the early inhibition of highly activated Tr1 cell development in first infection and the later amplification of Th1 and Tfh cell recall responses, particularly in second infection, suggesting that transient JAK/STAT inhibition functions not only as a direct immunomodulator but also as a microenvironmental stabiliser that promotes more durable anti-parasitic CD4⁺ T cell immunity.

CD4⁺ T cells exhibit developmental plasticity during infection, enabling them to adopt or transition between effector, regulatory, and helper states in response to dynamic inflammatory and tissue-derived cues (Gagliani and Huber, 2017; O’Shea and Paul, 2010). Therefore, Th1, Tfh, and Tr1 cells can share transcriptional programs, co-express lineage-defining molecules and interconvert depending on antigen load, cytokine exposure, metabolic stress, as well as the microanatomical niche in which they reside (Crotty, 2019). Inflammatory mediators such as type I interferons, IFNγ, IL-10, IL-12, and costimulatory or coinhibitory interactions can redirect differentiation trajectories, allowing CD4⁺ T cells to fine-tune their function as immune and tissue demands change (Masopust et al., 2025; Tuzlak et al., 2021). In malaria, this plasticity is well documented, and CD4⁺ T cells frequently display overlapping Th1, Tfh and Tr1 cell phenotypes, co-expressing IFN-γ, IL-10, and CXCR5, and shifting between inflammatory, regulatory, and B cell-helping roles as infection progresses (Boyle et al., 2024; Kurup et al., 2019). IL-10–producing Th1 cells generated during acute *P. falciparum* or *P. berghei* ANKA infection can acquire Tfh cell-like features and migrate into B cell follicles, while Tfh cells themselves may adopt Th1 or Tr1 cell characteristics under strong inflammatory pressure (Edwards et al., 2023; Obeng-Adjei et al., 2015; Ryg-Cornejo et al., 2016). Such plasticity likely helps balance parasite control with limiting immunopathology, though it may also compromise germinal centre responses in severe malaria (Boyle et al., 2024). Our results support this model by demonstrating plasticity within Th1, Tfh and Tr1 cells at both transcriptional and phenotypic levels. Rather than forming discrete, terminally differentiated subsets, these lineages shared transcriptional signatures, overlapping effector modules, and clonal relationships, consistent with dynamic interconversion during infection. This plasticity likely reflects adaptation to distinct microenvironmental cues encountered within specific tissue niches, particularly the spleen, where malaria-induced remodelling generates spatially and temporally heterogeneous cytokine and antigen landscapes (Engwerda et al., 2005; Kho et al., 2021). (Kumar et al., 2019; Wykes and Lewin, 2018)These findings suggest that the fate of anti-parasitic CD4⁺ T cell subsets is not pre-set but continuously moulded by their microenvironment, highlighting opportunities to modulate T cell states therapeutically to enhance protective immunity.

Previous studies on CD4^+^ T cells from Ugandan children living in a high, perennial malaria transmission region found Tr1 cells were the dominant CD4^+^ T cell subset after *P. falciparum* infection (Nideffer et al., 2025). These cells were capable of long-term, parasite-specific recall responses and scRNA/TCR sequencing indicated phenotypic clonal fidelity. In contrast, in our CHMI cohort we observe extensive TCR clonal sharing across Th1, Tfh and Tr1 cell subsets, with individual parasite-specific clones spanning different transcriptional and phenotypic states over time. Several biological and experimental factors likely account for these differences. First, Ugandan children and adults experience repeated natural infections over years, potentially generating highly imprinted, long-lived memory clones that appear to have stable Tr1 cell fates, whereas our malaria-naïve volunteers were sampled during primary and very early second infections after rapid parasite clearance, when CD4⁺ T cells are still transitioning between effector, B cell helping and regulatory states. Second, we deliberately perturbed type I IFN–JAK/STAT signalling with ruxolitinib, altering inflammatory cues and likely promoting dynamic repositioning of Th1, Tfh and Tr1 cell clones rather than reinforcing a single dominant Tr1 cell fate. Third, there are methodological differences: Nideffer *et al*. focused on relatively late, memory timepoints after substantial clonal expansion within well-defined memory subsets enabling a call of “clonal fidelity” (Nideffer et al., 2025), whereas our clone tracking included both *ex vivo* and AIM⁺ cells across acute and recall phases, capturing intermediate and mixed transcriptional states that would register as phenotypic heterogeneity rather than fidelity. Together, these distinctions suggest that high-fidelity Tr1 cell imprinting may be a hallmark of long-standing, naturally acquired malaria immunity in endemic settings, while in early infection and under targeted cytokine pathway modulation, malaria-specific CD4⁺ T cell clones remain highly plastic and redistribute across Th1, Tfh and Tr1 cell compartments.

Tfh cells play an important role in helping B cells produce parasite-specific antibodies necessary for effective parasite clearance (Boyle et al., 2024; Chan et al., 2022; Chan et al., 2020). Notably, in addition to promoting the expansion of Tr1 cells following a second infection (including those exhibiting Tfh cell characteristics), ruxolitinib treatment also led to expansion of Tfh1 cells, a subset critical for antibody production in viral infections such as HIV (Baiyegunhi et al., 2018), SARS-CoV-2 (Gong et al., 2020), and influenza (Bentebibel et al., 2013). Recent work has also shown that some subsets of Tfh1 cells associate with antibody development in controlled human malaria infection, together with Tfh2 cells (Soon et al., 2025). Ruxolitinib treatment also resulted in expansion of Tfh-Tr1 cells which shared phenotypes of both subsets. The implications of this shift in Tfh subset dynamics remain unclear, particularly as we found that despite improved control of parasite growth in all participants following a second infection, relative to the first infection, there was no ruxolitinib-mediated improvement (Webster et al., 2025). Nevertheless, changes to Tfh cell subsets may impact on the quality and quantity of the B cell and humoral response, and investigating these changes is the focus of ongoing research endeavours.

There are several limitations with our studies, including that it was conducted in malaria-naïve adults in Australia and treatment was administered relatively early during infection due to ethical constraints. Both these factors may have limited our ability to detect differences in parasite growth between participants treated with ruxolitinib and control, following second infection. Additionally, we were unable to establish how ruxolitinib would impact pre-existing anti-parasitic CD4^+^ T cells. Thus, a logical extension for this work would be to test ruxolitinib as a host-directed treatment in participants in an endemic setting, either during controlled human malaria or in natural infection, assessing influences on parasite control and development of immunity.

Together, these findings highlight the potential of transient JAK/STAT inhibition as a rational host-directed strategy to improve CD4⁺ T cell immunity during acute infection, balancing necessary suppression of early inflammation with enhanced generation of durable, high-quality anti-parasitic memory. By demonstrating that ruxolitinib can modulate CD4⁺ T cell signalling, delay regulatory skewing, potentially stabilise lymphoid microenvironments, and ultimately amplify recall responses by Th1, Tfh and Tr1 cells, our work provides early evidence for a strategy of deploying immune modulators to compliment other malaria control strategies. Importantly, these insights extend beyond malaria, offering new knowledge for how short-course cytokine pathway inhibition might be used to optimise vaccine- or drug-induced immunity in other infectious diseases characterised by acute inflammatory responses. Future studies in malaria endemic populations, incorporating longer-term clinical endpoints and evaluation of humoral and cellular protective correlates, will be essential to determine whether ruxolitinib or similar agents can safely enhance protective immunity in malaria endemic settings.

## Materials and Methods

### Ethics statement

Ethics approval for this study was obtained from the QIMRB Berghofer Human Research Ethics Committee (P3696 and P1479), and the Alfred Health Ethics Committee (288/23). Written informed consent was obtained from all study participants.

### Study cohort

Samples in this study were collected from a controlled human malaria infection cohort “A randomised, double blind, placebo-controlled trial to evaluate the safety, tolerability and anti-parasitic immunity boosting activation of ruxolitinib when co-administered with artemether-lumenfantrine in healthy volunteers with *Plasmodium falciparum* Induced Blood Stage Malaria”. The study is registered with the Australian New Zealand Clinical Trials Registry: ACTRN12621000866808. Study participants were healthy malaria-naïve adults aged 18 to 55 years who met the eligibility criteria (Webster et al., 2025).

Details of the study design are reported elsewhere (Webster et al., 2025). In brief, in the first phase of the study, 20 participants were intravenously inoculated with 3D7 *P. falciparum*-infected RBCs. Blood parasitemia was monitored from day 4 post-infection using qPCR, and on day 8 or 9 participants were randomised to receive artemether-lumefantrine (AL) with either ruxolitinib or placebo. Participants and investigators were blinded to treatment allocation. 15 participants returned for the second phase of the study and were re-inoculated with *P. falciparum*-infected RBCs 91 days post initial infection, and their blood parasitemia levels monitored as before. AL only was then administered once blood parasitemia levels reached or exceeded 50,000 parasites/ml. One of the participants contracted SARS-CoV-2 during the re-infection period and was therefore excluded from subsequent analyses.

Participants were monitored to ensure safety and parasite clearance up until six months post initial infection. Blood samples for immunological analyses were collected on designated days throughout the trial.

Whole blood from the study participants was collected into lithium heparin tubes and processed for whole blood assays or PBMCs were isolated with Ficoll-Paque (Sigma, USA), density gradient centrifugation. PBMCs were stored in cryopreservation media (90% (v/v) FBS, 10% (v/v) DMSO) in liquid nitrogen storage.

### Whole blood phospho-CYTOF

Whole blood samples were collected from the participants in the clinical trial at day 0 (baseline), at pre-treatment on day 8/9, on the second day of treatment day 9/10 (T+1.5, halfway through the treatment regimen) and day 13/14 (T+5, 5 days after the final treatment). Two hundred µl of whole blood (from lithium-heparin) was stimulated with 1µg/ml IFNβ or 50 ng/ml IL-2 or 50 ng/ml PMA and 0.4855 µg/ml Ionomycin, or left untreated for 15 min at room temperature, and then fixed with Proteomic Stabilizer (Smart Tube, Inc.) and kept at -80°C. CyTOF was performed by the Human Immune Monitoring Center (HIMC) at Stanford University. Fixed samples were thawed; samples were washed with Smart Tube Thaw-Lyse buffer twice and washed with Cell Staining Buffer (CSB; Standard BioTools) once. The samples where then washed twice with perm buffer (Invitrogen 10× permeabilization buffer #00-8333-56, diluted to 1× in phosphate-buffered saline [PBS] from Standard BioTools). The samples were then resuspended in 900 µl perm buffer and 10 µl of each Standard BioTools CellID 20plex Pd barcode was added before incubating at room temperature for 30 min. Next, the samples were centrifuged, aspirated, and washed three times by resuspension in CSB. The pellets were then resuspended in 450 µl CSB and pooled into a single tube. After centrifugation and aspiration, the pooled sample was counted on a Bio-Rad TC20 cell counter. Surface cocktail was added at 1 test, 20 µl per 1 million counted cells and stained at room temperature for 30 min (Supplementary Table S1). This cocktail was combined from two -80 °C frozen cocktails (HIMC surface stain and Boyle surface stain) to which fresh 143Nd-CD45RA (Standard BioTools) was added. Next, the sample was washed twice by resuspension with CSB. For fixation, 4% paraformaldehyde (PFA) in PBS was added and incubated for 10 min at room temperature before adding PBS to 10 ml, centrifugation, and aspiration. Then, 1 ml of -20°C cold methanol was added and the sample placed at -80°C overnight. The next day, the sample was centrifuged and the methanol aspirated before resuspension in 10 ml PBS and the centrifugation and aspiration repeated. The sample was resuspended in 1 ml CSB and counted by TC20 cell counter. Intracellular cocktail was added: 1 test, 20 µl per 1 million counted cells and stained at room temperature for 30 min. This cocktail was combined from two -80°C frozen cocktails (HIMC intracellular stain and Boyle intracellular stain). After staining, the sample was washed twice with CSB, centrifuged, and aspirated. The pellet was then resuspended in 2% PFA/PBS containing 250 nM Ir intercalator (Standard BioTools) and incubated for 20 min at room temperature. Cells were washed once with CSB and twice with MilliQ water. On the same day as staining was finished, cells were diluted to 750 000/ml in MilliQ water containing 10× diluted EQ4 normalization beads (Standard BioTools) and acquired by CyTOF, Helios, CyTOF model 3. Data analysis was performed using FlowJo v10 by gating on intact cells based on the iridium isotopes from the intercalator, then on singlets by Ir191 versus cell length followed by cell clustering analyses. No live-dead was used as the sample was fresh complete whole-blood. For clustering analyses, data was batch corrected using the cyCombine package (Pedersen et al., 2022), clustered using FlowSOM (Van Gassen et al., 2015), and annotated by expression profiles of lineage markers on R version 4.3.3. For CD4 T cell subset analysis, CD4 T cells were clustered with FlowSOM (Van Gassen et al., 2015), with CD45RA, CD27, CD38, HLA-DR, CD127 and CD25 and the default settings was used to subcluster the CD4^+^ T cells with a meta.k value of 15. The data was then subsampled to 5000 cells per participant and visualised using the run.umap function. A clustered heatmap and density plots showing the expression of the clustering markers were used to annotate the clusters. After identification of the CD4^+^ T cells, the create.sumtable function was used to obtain the frequency of the cells as a percentage of the total CD4^+^ T cells and the mean signal intensity (MSI) of the phosphorylated proteins on the cells for each stimulation condition.

### PBMC Immunophenotyping CyTOF

This assay was performed in the Human Immune Monitoring Center at Stanford University. PBMCs were thawed in warm RPMI+FBS media containing benzonase, washed twice, resuspended in CyFACS buffer (PBS supplemented with 2% BSA, 2 mM EDTA, and 0.1% sodium azide), and viable cells were counted by Vicell. Cells were added to a U-bottom microtiter plate at 1.5 million viable cells/well and washed once by pelleting and resuspension in fresh CyFACS buffer. The cells were stained for 60 min at room temperature with 50 μl of the following antibody-polymer conjugate cocktail (Supplementary Table S2). This cocktail was combined from two -80 °C frozen cocktails (HIMC surface stain and Boyle surface stain). All antibodies were from purified unconjugated, carrier-protein-free stocks from BD Biosciences, Biolegend, or R&D Systems. The polymer and metal isotopes were from Fluidigm Corporation. The cells were washed twice by pelleting and resuspension with 500 μl FACS buffer. The cells were resuspended in 100 μl PBS buffer containing 2 μg/ml Live-Dead (DOTA-maleimide (Macrocyclics) containing natural-abundance indium). The cells were washed twice by pelleting and resuspension with 500 μl PBS. The cells were resuspended in 100 μl 2% PFA in PBS and placed at 4° C overnight. The next day, 500 μl CyFACS was added, and the cells were pelleted. The cells were resuspended in 100 μl eBiosciences permeabilization buffer (1x in PBS) and placed on ice for 45 min before washing once with 500 μl CyFACS. The cells were resuspended in 100 μl iridium-containing DNA intercalator (1:2000 dilution in 1x PBS; Fluidigm) and incubated at room temperature for 20 min. The cells were washed once with 500 μl CyFACS and twice in 500 μl MilliQ water. The cells were diluted to 800 000/ml in MilliQ water containing a 10x dilution of EQ Normalization beads (Fluidigm) before injection into the CyTOF (Fluidigm). CD4^+^ T cells were identified within PBMC based on expression of all markers and then concatenated. Cells were then clustered according to the expression of the T cell markers (CD45RA, CCR7, CXCR3, CXCR5, CD127, CD25, LAG3 and CD49b) using FlowSOM clustering with the default Spectre settings. As a results of low LAG3 staining, an over-clustering approach was used with a meta.k value of 60 to identify cells expressing LAG3. The data was subsampled to 2,500 cells per sample and visualised using UMAP dimensionality reduction using the run.umap function default Spectre package settings. The 60 CD4^+^ T clusters were annotated according to the expression of the chemokine and activation markers and heatmap was created using the make.pheatmap function in Spectre. Following clustering, the create.sumtable function was used to generate a summary table of the frequencies of the cell subsets as a percent of the total CD4^+^ T cell population.

### PBMC, Tr1 and CD4^+^ T cell co-inhibitory receptor flow cytometry

Cryopreserved PBMCs were thawed in RPMI 1640 (Gibco, Life Technologies), containing 10% FBS (heat-inactivated) and 0.02% Benzonase Nuclease (Sigma-Aldrich), and thawed cells counted with a DeNovix Cell Drop FL. PBMCs (1x10^6^ per sample) were stained with antibodies against LAG3, CCR2, CCR5, TIGIT, and TIM3 (Pre-Stain, Supplementary Table 4) along with Human Fc block (Invitrogen, 1/30) in 2% FBS/RPMI (cRPMI) at 37 °C, 5% CO2 for 1 h. Cells were then stained with Surface Stain antibodies (Supplementary Table 3) in Brilliant Stain Buffer (1/10) diluted in 2% FBS/PBS at 37°C, 5% CO2 for a further 30 min. Cells were washed twice with PBS and stained with Live/Dead Blue (Invitrogen, 1/1000 in PBS) in the dark at room temperature for 15 min. Cells were washed twice with 2% FBS/PBS and fixed using FoxP3 eBioscience fixative/permeabilization buffer at 4°C in the dark for 30 min. Cells were washed twice with FoxP3 eBioscience Permeabilization buffer and stained with intracellular stain (Supplementary Table S3) in permeabilization buffer in the dark, at RT for 30 min. Cells were washed twice with permeabilization buffer and fixed with BD stabilising fixative incubated at room temperature for 10 min. Cells were washed and resuspended 2% FBS/PBS and acquired on the Cytek Aurora 5 flow cytometer. Data were analysed with FlowJo version 10.9.0. Dead cells and doublets were excluded and the CD3^+^ CD8^-^ CD4^+^ T cells were gated (Supplementary Figure 5). FCS files containing the CD4+ T cell data were exported and analysed in the R package Spectre (Ashhurst et al., 2022). Data was concatenated and cyCombine batch alignment was performed using the default cyCombine correct.batch.effects function. Tr1 cells were identified as NOT Tregs (CD127^low^ CD25^+^) and then as LAG3^+^/CD49b^+^ (Supplementary Figure 5). The Tr1 cells were clustered according to the expression of the T cell markers (CD45RA, CCR7, CXCR3, CXCR5, CCR2, CCR5, TIM3, TIGIT, CD38, ICOS, CTLA-4, PD-1, Ki67 and CD161) using FlowSOM clustering with meta.k value of 10 and the default Spectre settings. The Spectre make.pheatmap function was used to create a heat map to visualise the expression of the markers on the 10 Tr1 clusters and the create.sumtable function used to determine the frequencies of the clusters as a percentage of all Tr1 cells. The Tr1 cells were down sampled to 2,500 cells per sample and dimensionality reduction performed using the run.umap function.

### P. falciparum parasite culture

Packed red blood cells (RBCs, acquired from Australian Red Cross, Melbourne Australia) were infected with the *Plasmodium falciparum* 3D7 parasite strain used in CHMI (68) and cultured at 5% hematocrit in Roswell Park Memorial Institute 1640 media (RPMI) supplemented with AlbuMAX II (0.25%) and heat-inactivated human sera (5%), at 37 °C in 1% O2, 5% CO2, 94% N2 gas mixture. Culture media was replaced daily, and parasites monitored by Giemsa-stained blood smears. pRBCs were grown to 15% parasitemia and purified from uninfected RBCs (uRBCs) via magnet separation to enrich mature trophozoite stage pRBCs. Purified pRBCs (>95% purity) were stored at80 °C in Glycerolyte cryopreservant. For use in stimulations, glycerolyte preserved parasites or control uRBCs were thawed by dropwise addition of malaria thawing solution (MTS: 0.6% NaCl in H20) and incubated at room temperature for 10 min. Parasites were washed in MTS/PBS (phosphate bufferd saline) three times with decreasing solution ratios (1:0, 1:1, 0:1). Following thaw, parasites were intact late stage trophozoites and diluted to 1 x 10^6^ pRBC / 50µl.

### Activation Induced Marker (AIM) assay

PBMCs were thawed in RPMI 1640 (Gibco) containing 10% FCS and 0.02% Benzonase and rested for 2 hours at 37°C, 5% CO2, and then stimulated for 18 hours in 96-well plates in 10% FCS/RPMI at 1 x 10^6^ cells per well with 50 µl pRBCs or uRBCs (1 x 10^6^). Following stimulation, PBMCs were stained with antibodies to LAG3, CD49b and CCR7 (Pre-stain, Supplementary Table 4), in Human Fc block (2% FBS/RPMI) at 37°C, 5% CO2 for 45 min. Cells were washed and stained with Live/Dead Blue (Invitrogen, Thermo Fisher Scientific, Cat# L23015, 1/1,000 PBS) in the dark at room temperature for 15 min. Cells were washed twice with 2% FBS/PBS and stained with Surface Stain antibodies (Supplementary Table 4 in 2% FBS/PBS w room temperature for 15 min. Cells were washed twice with 2% FBS/PBS and fixed with BD Stabilizing fixative at room temperature for 20 min. Cells were then washed twice and resuspended in 2% FBS/PBS and acquired on Cytek Aurora 5 using Spectroflo software. Flow cytometry results were then analysed on FlowJo version 10.9.0. CD4^+^ T cell subsets identified based on FSC/SSC, Live, CD14^-^/CD19^-^/CD3^+^/CD4^+^ cells (Supplementary Figure 6), and AIM+ cells identified as CD69^+^/OX40^+^ and boolean gating was used to identify subsets (Supplementary Figure 6). Malaria specific AIM+ cells were calculated as % pRBC AIM+ CD4^+^ T cells minus % uRBC AIM+ CD4^+^ T cells. Negative values as a result of background subtraction were set to zero.

### Single Cell RNA sequencing and analysis

#### Cell isolation

PBMCs from four placebo and four ruxolitinib donors at ten timepoints (Supplementary Table 5) were thawed in RPMI 1640 (Gibco) containing 10% FCS and 0.02% Benzonase. Cells were rested for 2 h in 10% FCS/PBS at 37 °C, 5% CO2 and then incubated with Fc block for 15 min and surface stained with antibodies listed in Supplementary Table 6 for 30 min at 4°C. Cells were washed with 1% BSA/PBS and stained for viability with Sytox Blue (Invitrogen (CA, USA), Cat. # S34857) and sorted on BD FACSAria™III Cell Sorter into 1% BSA/PBS. For AIM^+^ CD4^+^ T cells, PBMCS were first stimulated with pRBCs as described for AIM assays, then processed as above for unstimulated samples. AIM^+^ CD4^+^ T cells were identified as CD4^+^CD3^+^CD45RA^-^OX40^+^CD69^+^ cells. As AIM assays were performed across multiple days (two donors per experiment) for logistical reasons, a control PBMC sample (HMI16) was included in each assay to rule out potential batch effects across assays.

### 10 X Genomics Chromium GEX Library preparation and sequencing

Sorted cells were loaded into each lane of Chromium Next GEM Single Cell 5’ Reagent Kit v2 (Ex vivo: 6-7 samples were multiplexed per lane; AIM^+^: 17-19 samples were multiplexed per lane) and Gel Bead-in- Emulsion (GEMs) generated in the 10X Chromium Controller (Chromium Next GEM Single Cell 5’ Kit v2, PN-1000263, Chromium Single cell V(D)J Amplification Kits: Human TCR, PN-1000252, Chromium Next GEM Chip K Single Cell Kit, PN-1000286, Dual Index Kit TT Set A, PN-1000215). 5’ Gene Expression Libraries and VDJ libraries (TCR) were generated according to the manufacturer’s instructions. Generated libraries were sequenced in an Illumina NextSeq 2000 System using P3 flow cells according to the manufacturer’s protocol using paired-end sequencing with the following parameters Read 1: 26 cycles, i7 index: 10 cycles, i5 index: 10 cycles and Read 2: 90 cycles.

#### Demultiplexing and analysis

Each scRNAseq data set was demultiplexed, aligned and quantified using Cell Ranger software (version 6.0.1;10x Genomics) against the human reference genome (GRCh38-3.0.0) with default parameters. Cells were first demultiplexed using the *HTODemux()* function in Seurat based on sample hashtag oligo (HTO) information. To further confirm donor identity, genotype-based demultiplexing was performed using *cell-snp lite* (version 1.2.2) (Huang and Huang, 2021) to call SNPs from 1000_Genome_Project (Auton et al., 2015) reference panel, followed by *Vireo* (version 0.2.0), in Python (version 3.6.1), to assign each cell to the original donor based on haplotype ratios (Huang et al., 2019). Only cells predicted as singlets were retained for downstream analysis. Cell ranger count matrices for each sample were loaded, merged and analysed using the Seurat package (version 5.1.0) (Hao et al., 2024). Cells were filtered to remove those expressing fewer than 200 genes and more than 6000 genes, and those with more than 5% mitochondrial content. Only genes expressed in three or more cells were considered. TRA/TRB gene expression were removed from the GEX dataset, as well as a small population of contaminating CD8^+^ T cells and monocytes, prior to further downstream processing.

Dataset was split by donor and then each individual donor dataset was normalised by ‘SCTransform.v2’ with regression of mitochondrial mapping percentage. Harmony integration was then performed using the *IntegrateLayers()* function to correct for inter-donor batch effects. Principal component analysis (PCA) was performed using *RunPCA()* and the top 30 principal components were used to compute UMAP visualisation using *RunUMAP()*.

#### Cell cluster annotation

Clustering and cell annotation were performed separately for the *ex vivo* and AIM datasets. The integrated dataset was first divided by assay (*ex vivo* and AIM), and each dataset was processed independently by performing SCTransform normalisation, Harmony integration by donor, PCA dimensionality reduction and UMAP visualisation as described above. The AIM dataset included samples from only six donors, as two donors were excluded due to technical limitations. *FindNeighbors()* was run using the top 30 principal components and *FindClusters()* was used to perform unsupervised clustering at a resolution of 1- 1.2 (*ex vivo* dataset) or 0.5 (AIM dataset). Cluster marker genes were identified using *FindMarkers()* and clusters were annotated and merged based on canonical cluster marker expression, with ten and eight cell subsets identified for the *ex vivo* and AIM datasets, respectively. The Upset plot was generated using the UpSetR package (Conway et al., 2017) (version 1.4.0) to look at intersections of differentially expressed cluster marker genes between CD4^+^ T cell subsets.

#### Gene signature scoring

The Seurat *AddModuleScore()* function was used to calculate gene signatures.

#### Pseudobulked Differential Gene Expression Analysis and Pathway Analysis

For each cluster (Tr1, Tfh, Th1) and timepoint (T+7, rT+3) raw count data and metadata were extracted from the Seurat objects. Counts were aggregated per donor to generate pseudobulk expression profiles for each cluster and timepoint. Differential expression analysis was performed using edgeR (version 4.6.2) (Chen et al., 2025). Raw counts were supplied to *DGEList*, and normalised using the TMM method (*calcNormFactors*), and dispersions estimated. Quasi-likelihood (QL) F-tests were performed using *glmQLFit* and *glmQLFTest* to identify genes differentially expressed between ruxolitinib- and placebo-treated groups. Genes with P < 0.05 were considered significant. Volcano plots were generated using *ggplot2* (version 3.5.1) and *ggrepel* (version 0.9.6). Pathway enrichment analysis was performed for each cluster and timepoint using the *SCPA* package (Bibby et al., 2022) (version 1.6.2), based on the Hallmark gene sets from MSigDB (Liberzon et al., 2015).

#### TCRA/B clonal analysis

Annotated TCR sequences were integrated with the single-cell transcriptomics data using the *combineTCR()* and *combineExpression()* functions from scRepertoire (version 2.5.0) (Yang et al., 2025). The CDR3 amino acid sequences were used to call and quantify clones. Only cells with single and complete paired TCRA and TCRB chain were included. Clones were defined as cells sharing identical amino acid sequence in both TCRA and TCRB. Malaria-specific clonotypes were defined as expanded clones (≥2 cells per clonotype) identified within the AIM^+^ dataset. These paired TCRA/B clonotypes were cross-referenced against the *ex-vivo* CD4^+^ T cell data by exact matching of the CDR3 amino acid sequences. Clonal overlap between samples was assessed using the *clonalOverlap()* function in scRepertoire, with similarity calculated using Jaccard’s index and visualised on heatmaps.

### Statistical analysis

The change in CD4 T cell responses over time from either the inoculation or the treatment timepoint was assessed using log-linear mixed effects models with restricted maximum likelihood (REML) estimation, incorporating a random intercept at the participant level. Prior to log10 transformation and modelling, data were transformed by adding half of the lowest value to the zero values. For changes between the first inoculation and treatment timepoint, only timepoint was added as a fixed effect. Treatment, timepoint, and their interaction were included as fixed effects to evaluate how the response differs between treatment groups and how the change in response over time differs between treatment groups. The change over time for the ruxolitinib group was assessed using contrasts. Residual bootstrapping for linear mixed effects models at 5000 iterations was applied to account for potential deviations from normality in the residuals, ensuring robust and reliable estimates. The analysis was performed in R using R Studio (version 4.3.1) using the Linear and Nonlinear mixed effects model (nlme) package (version 3.1-163) and the Fit linear mixed-effects models’ package lme4 (version 1.1-35-5). Non-normal residues were addressed using residual bootstrapping with the boot (version 1.3-30) and the lmeresampler packages (version 0.2.4). All plots were made with the R package ggplot2 (Wilkinson, 2011) or ggpubr (version 0.6.0) (Kassambara, 2018).

### Data and code availability

The raw sequencing data and processed Cell Ranger outputs used in this study have been deposited in the European Genome Archive data base under accession code EGAS50000001569.

Code used to analyse scRNAseq data is available at DOI: 10.5281/zenodo.18263999.

The processed single cell Seurat objects and flow cytometry files for AIM+ CD4^+^ T cells and Tr1 CD4^+^ T cells are available at OSF: DOI 10.17605/OSF.IO/JE7YD.

The pCyTOF whole blood data and PBMC CyTOF data are publicly available at: DOI 10.5281/zenodo.14872946 and 10.5281/zenodo.14873240.

## Data Availability

The raw sequencing data and processed Cell Ranger outputs used in this study have been deposited in the European Genome Archive data base under accession code EGAS50000001569.
Code used to analyse scRNAseq data is available at DOI: 10.5281/zenodo.18263999.
The processed single cell Seurat objects and flow cytometry files for AIM+ CD4+ T cells and Tr1 CD4+ T cells are available at OSF: DOI 10.17605/OSF.IO/JE7YD.
The pCyTOF whole blood data and PBMC CyTOF data are publicly available at: DOI 10.5281/zenodo.14872946 and 10.5281/zenodo.14873240.

## Author contributions

TW, SC, JJM, JSM, BEB, MJB and CRE conceptualised study

LB, JAE, DO FLR, MS, DA, ND, MN, RM, TMF, JH, JL and ML performed experiments and analysed data.

MN, FLR, JE, LB optimised protocols

BEB led the clinical trial.

RW provided project management for the clinical trial.

CRE, MJB, BEB, HM, GO provided supervision

RW, FA, GO and HM managed the logistics of sample collection and processing.

MJB, CRE, BEB, JSM, TW, SC, JJM, JSM provided funding

TW, SC, JJM, JSM, BEB, MJB and CRE conceived and managed the project, analysed the data and wrote the manuscript with feedback from all authors.

## Acknowledgments

QIMR-Berghofer and Burnet Institute acknowledge the traditional custodians of the lands where they are located, the Turrbal and Jagera people of Meanjin, and the Boonwurrung and Wurundjeri Woi-wurrong people of the Kulin Nation, respectively.

RBC and human serum were provided by the Australian Red Cross Blood Bank (Melbourne and Brisbane). We thank the volunteers who participated in the clinical trial; staff at the University of Sunshine Coast clinical trial site; staff at the Queensland Paediatric Infectious Diseases Laboratory (QPID) for conducting the malaria PCR assays; members of the Stanford Human Immune Monitoring Center for technical assistance;

This study was funded by the Australian National Health and Medical Research Council (NHMRC) Ideas Grant to BEB and MJB (GNT2002957). BEB and JSM are supported by NHMRC Investigator Grants (2016792 and 2016396, respectively); MJB is supported by a Snow Medical Fellowship (2022/SF167) and CSL Centenary Fellowship.

Work in the Stanford Human Immune Monitoring Center was supported by the Gates Foundation [INV-008378]. The conclusions and opinions expressed in this work are those of the author(s) alone and shall not be attributed to the Foundation. Under the grant conditions of the Gates Foundation, a Creative Commons Attribution 4.0 License has been assigned to the Author Accepted version of the manuscript.

## Supplementary Tables

**Supplementary Table S1.**
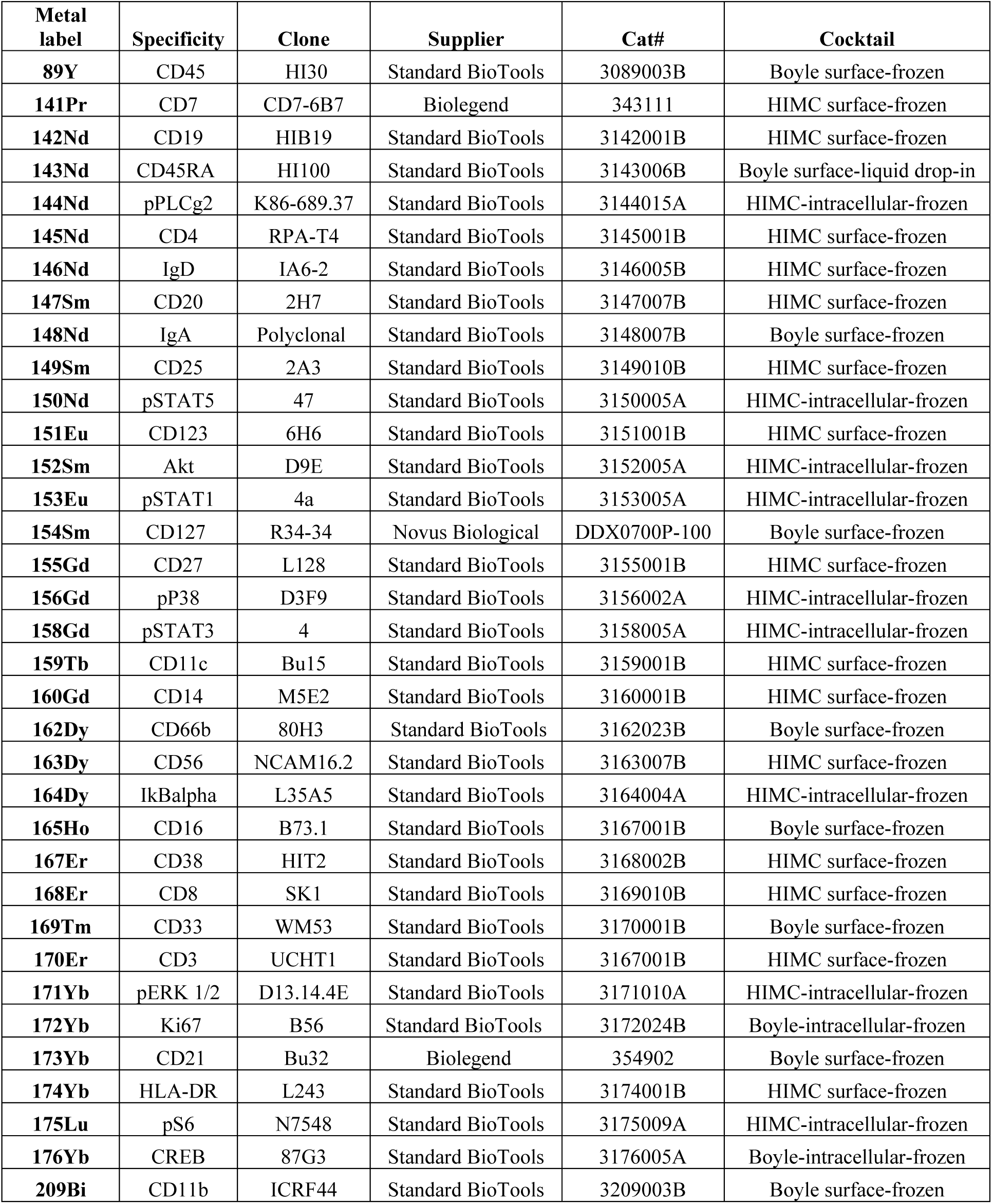
pCyTOF whole blood staining panel.

**Supplementary Table S2:** PBMC immunophenotyping CyTOF panel.

| **Metal label** | **Specificity** | **Clone** | **Supplier** | **Cat#** | **Cocktail** |
| --- | --- | --- | --- | --- | --- |
| **113In** | CD57 | HNK-1 | Biologend | 359602 | HIMC frozen |
| **115In** | live/dead |  |  |  |  |
| **141Pr** | CCR6 | G034E3 | Standard BioTools | 3141003A | Boyle frozen |
| **142Nd** | CD19 | SJ25C1 | Biologend | 363002 | HIMC frozen |
| **143Nd** | CD4 | SK3 | Biologend | 344602 | HIMC frozen |
| **144Nd** | CD8 | SK1 | Biologend | 344702 | HIMC frozen |
| **145Nd** | CD39 | A1 | Biologend | 328202 | HIMC frozen |
| **146Nd** | IgD | IA6-2 | Biologend | 348202 | HIMC frozen |
| **147Sm** | CD85j | 292319 | R&D Systems | MAB20172 | HIMC frozen |
| **148Nd** | CD11c | Bu15 | Biologend | 337202 | HIMC frozen |
| **149Sm** | CD16 | 3G8 | Biologend | 302002 | HIMC frozen |
| **150Nd** | CD3 | UCHT1 | Biologend | 300402 | HIMC frozen |
| **151Eu** | CD38 | HB-7 | Biologend | 356602 | HIMC frozen |
| **152Sm** | CD27 | L128 | BD Biosciences | special order | HIMC frozen |
| **153Eu** | CD21 | Bu32 | Biologend | 354902 | Boyle frozen |
| **154Sm** | CD14 | M5E2 | Biologend | 301802 | HIMC frozen |
| **155Gd** | Vd2 TCR | B6 | Biologend | 331402 | Boyle frozen |
| **156Gd** | CD94 | HP-3D9 | BD Biosciences | 555887 | HIMC frozen |
| **157Gd** | CD86 | IT2.2 | Biologend | 305402 | HIMC frozen |
| **158Gd** | CXCR5 | RF8B2 | BD Biosciences | 552032 | HIMC frozen |
| **159Tb** | CXCR3 | G025H7 | Biologend | 353702 | HIMC frozen |
| **160Gd** | CCR7 | 150503 | R&D Systems | MAB197 | HIMC frozen |
| **162Dy** | CD45RA | HI100 | Biologend | 304102 | HIMC frozen |
| **164Dy** | CD20 | 2H7 | Biologend | 302302 | HIMC frozen |
| **165Ho** | CD127 | A019D5 | Biologend | 351302 | HIMC frozen |
| **166Er** | CD33 | P67.6 | Biologend | 366602 | HIMC frozen |
| **167Er** | LAG3 | 11C3C65 | Biologend | 369302 | Boyle frozen |
| **168Er** | CD49b | P1E6-C5 | Biologend | 359302 | Boyle frozen |
| **169Tm** | ICOS | DX29 | BD Biosciences | 557801 | HIMC frozen |
| **170Er** | CD161 | DX12 | BD Biosciences | 556079 | HIMC frozen |
| **171Yb** | TCRgd | B1 | Biologend | 331202 | HIMC frozen |
| **172Yb** | PD-1 | EH12.1 | BD Biosciences | 562138 | HIMC frozen |
| **173Yb** | CD123 | 9F5 | BD Biosciences | 555642 | HIMC frozen |
| **174Yb** | CD56 | NCAM16.2 | BD Biosciences | 559043 | HIMC frozen |
| **175Lu** | HLA-DR | G46-6 | BD Biosciences | 556642 | HIMC frozen |
| **176Yb** | CD25 | M-A251 | Biologend | 356102 | HIMC frozen |
| **209Bi** | CD11b | ICRF44 | Standard BioTools | 3209003B | Boyle frozen |

**Supplementary Table 3:** PBMC CD4 T cells, Tr1 coinhibitory flow cytometry panel.

| Target | Fluorophor | Clone | Source | Dilution | Catalogue # |
| --- | --- | --- | --- | --- | --- |
| *Pre-Stain* |  |  |  |  |  |
| LAG3 | Alexa Fluor 647 | 11C3C65 | Biolegend | 1/50 | 369304 |
| CCR2 | APC-Cy7 | K036C2 | Biolegend | 1/120 | 357220 |
| CCR5 | BV 650 | 3A9 | BD Biosciences | 1/80 | 564999 |
| TIGIT | PE Dazzle | VSTM3 | Biolegend |  | 372716 |
| TIM3 | BV605 | F38-2E2 | Thermo Fisher | 1/100 | 16-3109-85 |
| *Surface Stain* |  |  |  |  |  |
| CD161 | Alexa Fluor 700 | HP-3G10 | Biolegend | 1/200 | 339942 |
| CD45RA | BUV 563 | HI100 | BD Biosciences | 1/800 | 565702 |
| CD4 | BUV 737 | SK3 | BD Biosciences | 1/120 | 564305 |
| CD8 | BUV 496 | RPA-T8 | BD Biosciences | 1/800 | 612942 |
| CD3 | BUV 805 | SK7 | BD Biosciences | 1/500 | 612893 |
| CXCR3 | BV421 | IC6/CXCR3 | BD Biosciences | 1/100 | 562558 |
| CD38 | BV480 | HIT2 | BD Biosciences | 1/50 | 566137 |
| CD14 | BV510 | M5E2 | Biolegend | 1/100 | 301842 |
| CD19 | BV510 | SJ25C1 | Biolegend | 1/100 | 363020 |
| CD127 | BV570 | A019D5 | Biolegend | 1/50 | 351308 |
| CXCR5 | BV711 | J252D4 | Biolegend | 1/50 | 356934 |
| ICOS | BV750 | C398.4A | Biolegend | 1/100 | 313558 |
| CD49b | FITC | Y418 | Invitrogen | 1/100 | 11-0498-42 |
| PD1 | PE/Cy7 | EH12.1 | BD Biosciences | 1/100 | 561272 |
| CD16 | PE/Fire 640 | 3G8 | Biolegend | 1/200 | 302068 |
| CD25 | PE/Fire 700 | M-A251 | Biolegend | 1/100 | 356146 |
| CCR7 | PerCP Cy5.5 | 150503 | BD Biosciences | 1/80 | 561144 |
| *Intra-cellular Stain* |  |  |  |  |  |
| CTLA-4 | BV750 | BNI3 | BD Biosciences | 1/120 | 563931 |
| Ki67 | BUV395 | B56 | BD Biosciences | 1/200 | 564071 |
| NKG7 | PE | 2G9A10F5 | Beckman Coulter | 1/2000 | IM3293 |

**Supplementary Table 4:** Antigen-induced marker assay flow cytometry panel.

| **Antibody** |  | **Clone** | **Source** | **Dilution** | **Catalogue Number** |
| --- | --- | --- | --- | --- | --- |
| *Pre-Stain* |  |  |  |  |  |
| LAG3 | BV421 | T47-530 | BD Biosciences | 1/50 | 565720 |
| CD49b | FITC | Y418 | Invitrogen | 1/100 | 11-0498-42 |
| CCR7 | PerCP Cy5.5 | G043H7 | Biolegend | 1/25 | 353220 |
| *Surface Stain* |  |  |  |  |  |
| CD161 | AF700 | HP-3G10 | Biolegend | 1/50 | 339942 |
| OX40 | APC | ACT35 | BD Biosciences | 1/50 | 563473 |
| CD45RA | BUV563 | HI100 | BD Biosciences | 1/400 | 565702 |
| CD8 | BUV496 | RPA-T8 | BD Biosciences | 1/100 | 612942 |
| CD3 | BUV805 | SK7 | BD Biosciences | 1/50 | 612893 |
| CD69 | BUV737 | FN50 | BD Biosciences | 1/100 | 612817 |
| CD14 | BV510 | M5E2 | Biolegend | 1/100 | 301842 |
| CD19 | BV510 | HIB19 | Biolegend | 1/100 | 302241 |
| CCR4 | BV605 | L291H4 | Biolegend | 1/100 | 359418 |
| CCR6 | BV650 | 11A9 | BD Biosciences | 1/50 | 563922 |
| CXCR5 | BV711 | J252D4 | Biolegend | 1/50 | 356934 |
| CD4 | BV785 | OKT4 | Biolegend | 1/50 | 317442 |
| CD25 | PE | BC96 | Biolegend | 1/200 | 302906 |
| PD1 | PE/Cy7 | EH12.1 | BD Biosciences | 1/50 | 561272 |

**Supplementary Table 5:**
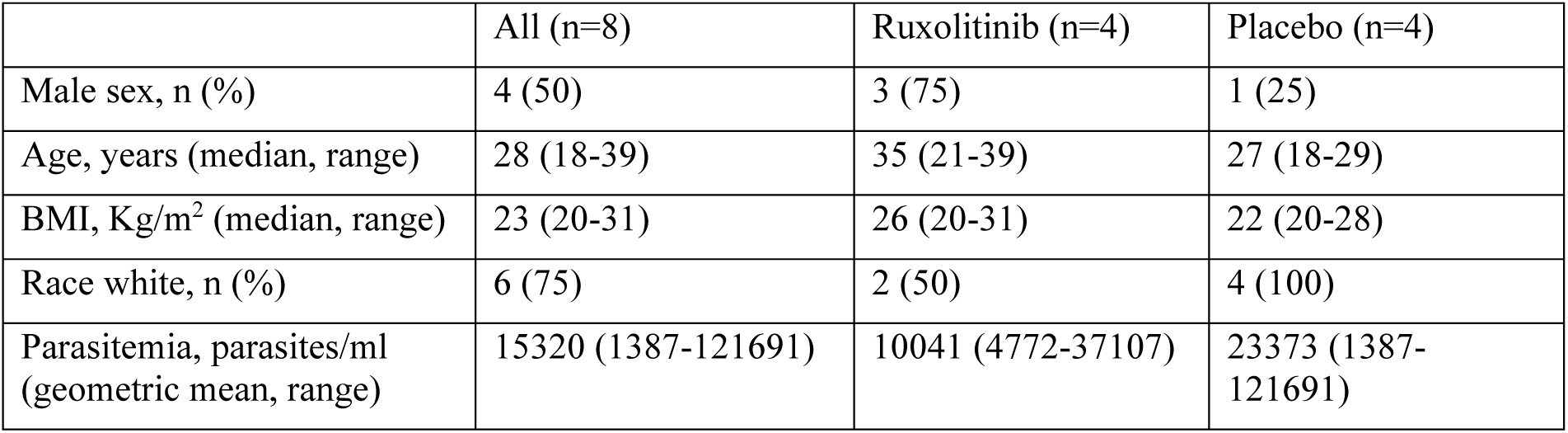
scRNAseq participant characteristics and parasitemia at time of ruxolitinib or placebo administration. BMI, body mass index.

**Supplementary Table 6:** Antibodies used to stain cells for scRNAseq.

| Antibody |  | Clone | Source | Dilution | Catalogue Number |
| --- | --- | --- | --- | --- | --- |
| *Surface Stain* |  |  |  |  |  |
| *Ex vivo dataset:* |  |  |  |  |  |
| CD3 | BV510 | SK7 | Biolegend | 1/50 | 344828 |
| CD4 | AF647 | SK3 | Biolegend | 1/100 | 344636 |
| CD45RA | BB515 | HI100 | BD Biosciences | 1/400 | 564552 |
| CD19 | PE | H1B19 | Biolegend | 1/100 | 302208 |
| *AIM dataset:* |  |  |  |  |  |
| CD3 | AF700 | SK7 | Biolegend | 1/50 | 344822 |
| CD4 | PerCP Cy5.5 | OKT4 | Biolegend | 1/50 | 317428 |
| CD69 | PE | FN50 | Biolegend | 1/50 | 310906 |
| OX40 | APC | ACT35 | BD Biosciences | 1/50 | 563473 |
| *Hashtags* |  |  |  |  |  |
| TotalSeq-C0251 anti-human Hashtag 1 |  | LNH-94; 2M2 | Biolegend | 0.3ug/sample | 394661 |
| TotalSeq-C0252 anti-human Hashtag 2 |  | LNH-94; 2M2 | Biolegend | 0.3ug/sample | 394663 |
| TotalSeq-C0253 anti-human Hashtag 3 |  | LNH-94; 2M2 | Biolegend | 0.3ug/sample | 394665 |
| TotalSeq-C0254 anti-human Hashtag 4 |  | LNH-94; 2M2 | Biolegend | 0.3ug/sample | 394667 |
| TotalSeq-C0255 anti-human Hashtag 5 |  | LNH-94; 2M2 | Biolegend | 0.3ug/sample | 394669 |
| TotalSeq-C0256 anti-human Hashtag 6 |  | LNH-94; 2M2 | Biolegend | 0.3ug/sample | 394671 |
| TotalSeq-C0257 anti-human Hashtag 7 |  | LNH-94; 2M2 | Biolegend | 0.3ug/sample | 394673 |
| TotalSeq-C0258 anti-human Hashtag 8 |  | LNH-94; 2M2 | Biolegend | 0.3ug/sample | 394675 |
| TotalSeq-C0259 anti-human Hashtag 9 |  | LNH-94; 2M2 | Biolegend | 0.3ug/sample | 394677 |
| TotalSeq-C0260 anti-human Hashtag 10 |  | LNH-94; 2M2 | Biolegend | 0.3ug/sample | 394679 |
| TotalSeq-C0296 anti-human Hashtag 11 |  | LNH-94; 2M2 | Biolegend | 0.3ug/sample | 328941 |
| TotalSeq-C0262 anti-human Hashtag 12 |  | LNH-94; 2M2 | Biolegend | 0.3ug/sample | 394683 |
| TotalSeq-C0263 anti-human Hashtag 13 |  | LNH-94; 2M2 | Biolegend | 0.3ug/sample | 394685 |
| TotalSeq-C0264 anti-human Hashtag 14 |  | LNH-94; 2M2 | Biolegend | 0.3ug/sample | 394687 |
| TotalSeq-C0265 anti-human Hashtag 15 |  | LNH-94; 2M2 | Biolegend | 0.3ug/sample | 394689 |
| TotalSeq-C0276 anti-human Hashtag 16 |  | LNH-94; 2M2 | Biolegend | 0.3ug/sample | 394691 |
| TotalSeq-C0277 anti-human Hashtag 17 |  | LNH-94; 2M2 | Biolegend | 0.3ug/sample | 394621 |
| TotalSeq-C0277 anti-human Hashtag 18 |  | LNH-94; 2M2 | Biolegend | 0.3ug/sample | 394693 |
| TotalSeq-C0279 anti-human Hashtag 19 |  | LNH-94; 2M2 | Biolegend | 0.3ug/sample | 394695 |

## Supplementary Figures

**Supplementary Figure 1:**
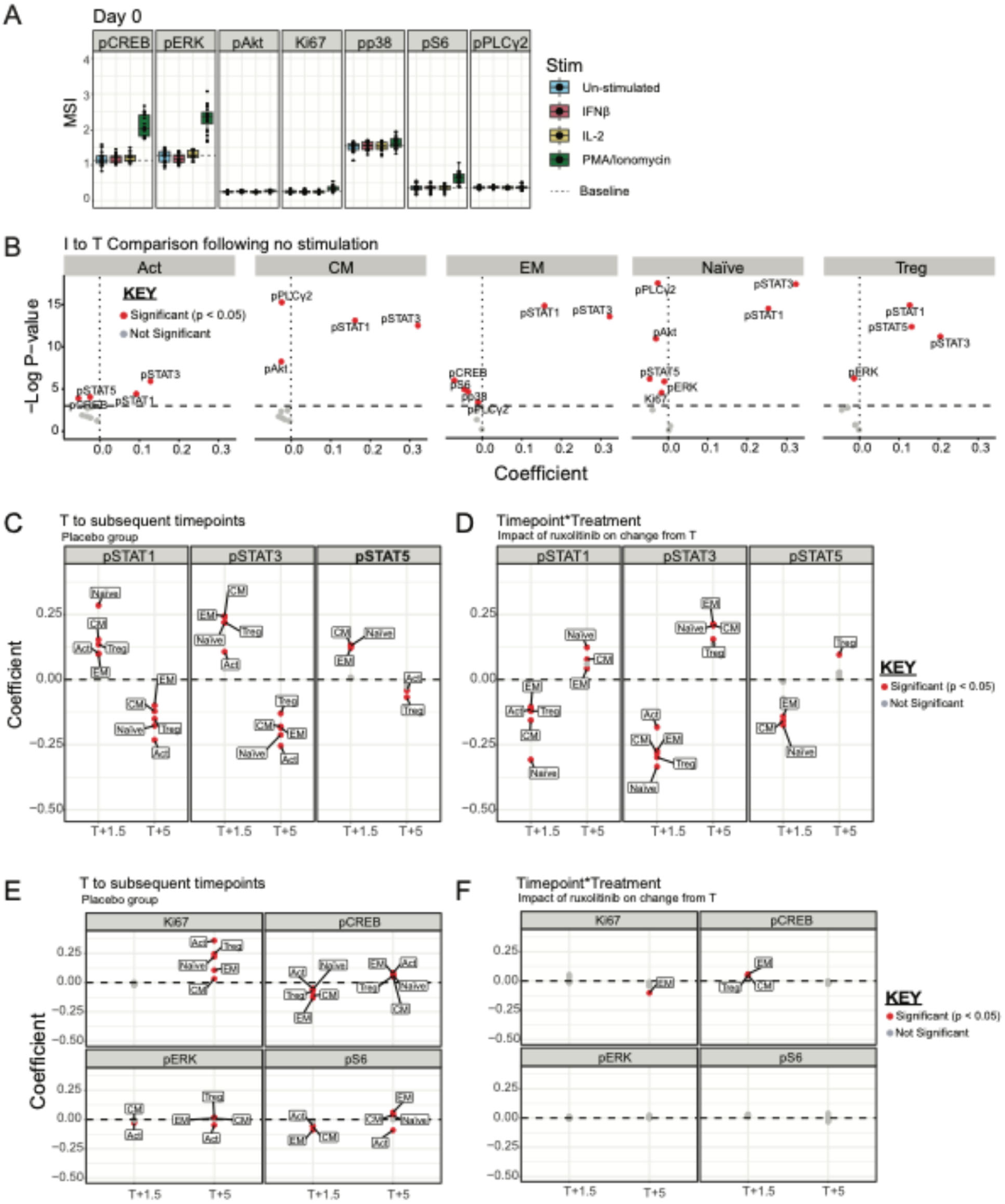
Analysis of phosphoproteins in CD4 T cells during malaria. **A**. Mean Signal Intensity (MSI) of phosphorylated proteins in CD4+ T cells following stimulation of whole blood with PBS (un-stimulated condition), IFNβ, IL-2 and PMA/Ionomycin at the pre-inoculation (baseline) time point for 15 minutes. The box plots show the extent of the lower and upper quartiles and the median, while each point represents a study participant (n=20). **B.** Volcano Plots showing the coefficient and p values of mixed model analysis of the effect of infection (pre-treatment time point (T, day 8/9) compared to inoculation time point (I, day 0)), on the mean signal intensity (MSI) of the phosphorylated proteins and Ki67 in CD4+ T cell subsets in unstimulated cells. Proteins with a significant change are in red. **C-F.** Coefficient from mixed effect models analysing MSI for phosphoproteins in unstimulated cells in indicated CD4 T cell subset, analysing changes with time (**C/E)**, comparing T with T+1.5 and T+5 and for the interaction between timepoint and treatment **(D/F**). Coefficient is indicated for each subset and timepoint, with significant changes in red. Related to Figure 1

**Supplementary Figure 2:**
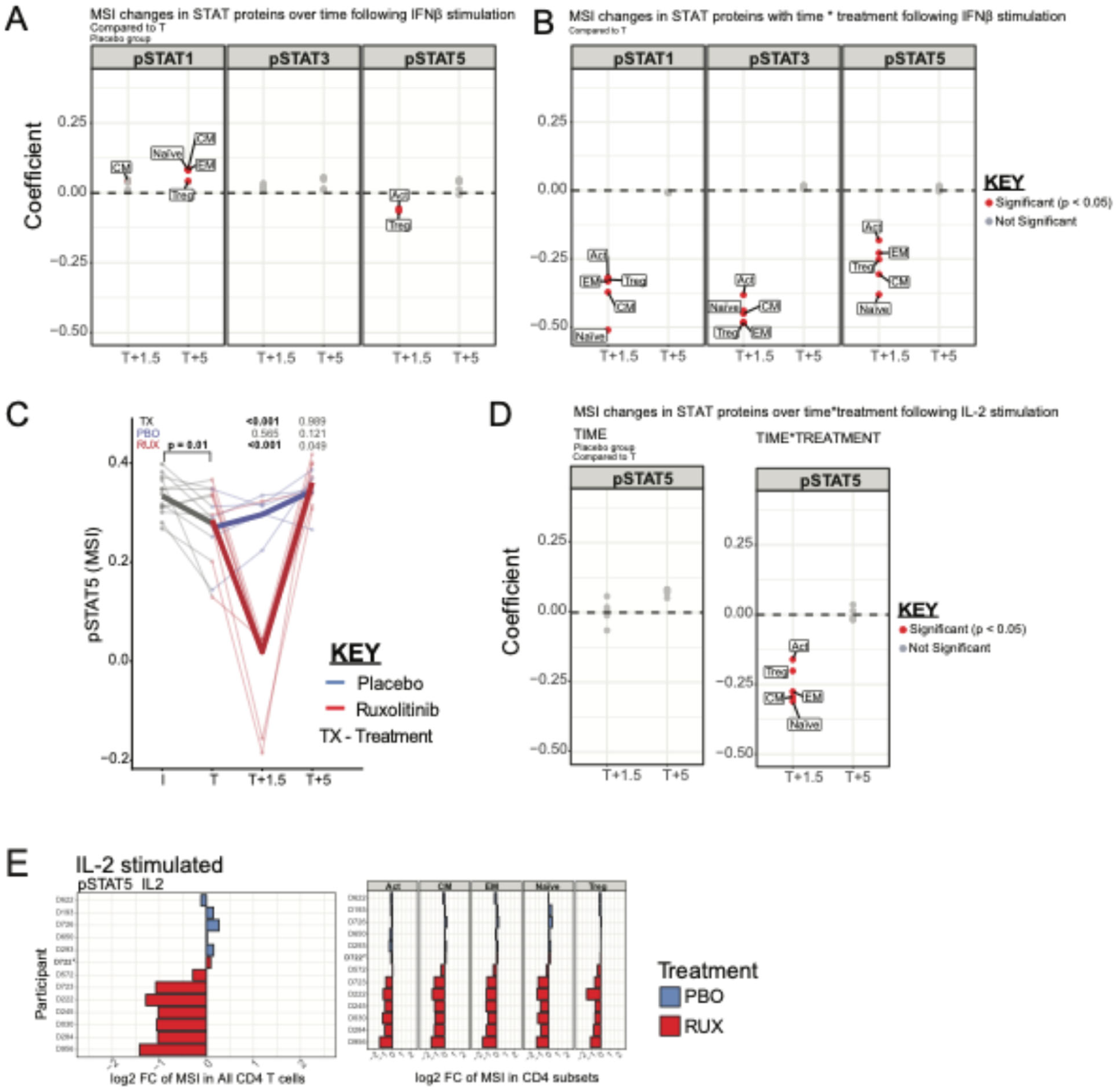
pSTATs in CD4 T cells during malaria following IFNβ and IL-2 stimulations. **A)** Coefficient from mixed effect models analysing MSI for phosphoproteins in cells stimulated with IFNb in indicated CD4 T cell subset, analysing changes with time (**A)** comparing T with T+1.5 and T+5 and for the interaction between timepoint and treatment **(B**). Coefficient is indicated for each subset and timepoint, with significant changes in red. **C)** pSTAT5 expression in CD4 T cells at I, T, T+1, T+5 in IL-2 stimulated cultures. Data are log transformed median signalling intensity, with thin lines representing each participant and coloured by treatment group. Prior to randomisation data are in grey; following randomisation ruxolitinib-treated participants are red and placebo-treated are blue. TX is p values for the interaction term between each timepoint (compared to T) and treatment groups. p values for the comparison between each timepoint and T are shown for the placebo (PBO) and ruxolitinib (RUX) groups, and were determined from contrasts. **D)** Coefficient from mixed effect models analysing MSI for pSTAT5 in cells stimulated with IL-2 in indicated CD4 T cell subset, analysing changes with time comparing T with T+1.5 and T+5 and for the interaction between timepoint and treatment. Significant changes are indicated in red. **E)** Fold change between T and T+1.5 expression of pSTAT5 in total CD4 T cells and CD4 T cell subsets for IL-2 stimulated cells. Data for each participant are shown. Related to Figure 1

**Supplementary Figure 3:**
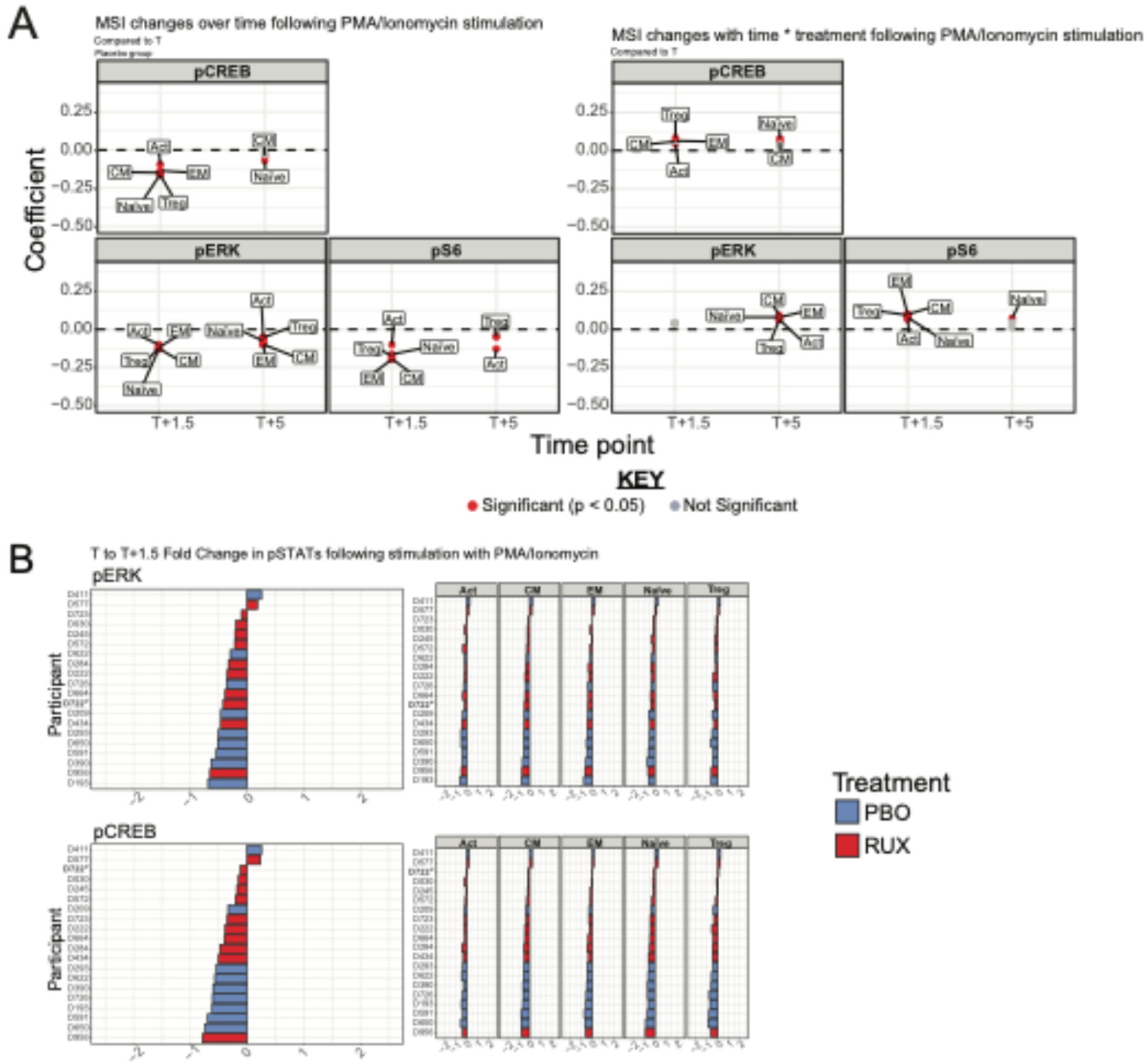
Phosphorylation of proteins in PMA stimulated cultures in CD4 T cells during malaria. **A)** Coefficient from mixed effect models analysing MSI for pCREB, pERK and pS6 in cells stimulated with PMA/Ion in indicated CD4 T cell subset, analysing changes with time comparing T with T+1.5 and T+5 and for the interaction between timepoint and treatment. Significant changes are indicated in red. **B)** Fold change between T and T+1.5 expression of pERK and pCREB in total CD4 T cells and CD4 T cell subsets for PMA/Ion stimulated cells. Data for each participant are shown. Related to Figure 1

**Supplementary Figure 4:**
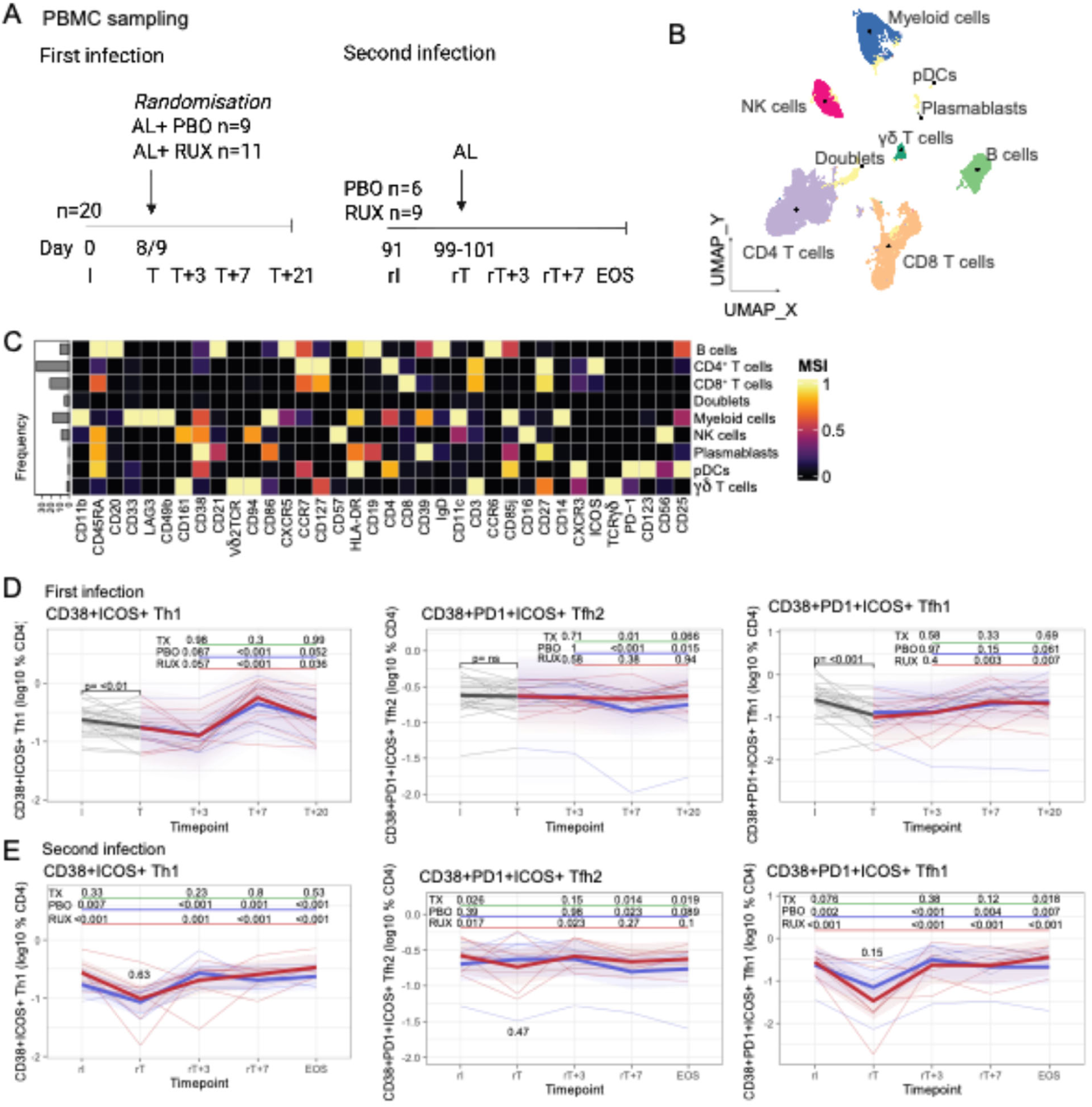
CD4 T cell frequencies and activation. **A)** PBMCs were collected at 5 time points in first and second infection. *one participant in the Rux group in second infection tested positive for COVID-19 and received malaria treatment early. **B/C)** PBMCs were analysed by CyTOF and CD4 T cells identified for further analysis. **D/E)** CD38+ICOS+ Th1, and CD38+PD1+ICOS+ Tfh2 and Tfh1 CD4 T cells in first and second infection. Data are log10 frequencies of CD4 T cells, with thin lines representing each participant. Prior to randomisation data are in grey, following randomisation ruxolitinib-treated participants are in red and placebo-treated are in blue. P values are from linear mixed effect models, with bold lines representing the mean of the predicted values from the fitted models for each group. TX is p values for the interaction term between each timepoint (compared to T) and treatment groups (underlined in green). P values for the comparison between each timepoint and T are shown for the placebo (PBO, underlined in blue) and ruxolitinib (RUX, underlined in red) groups, and were determined from contrasts. Related to Figure 2.

**Supplementary Figure 5:**
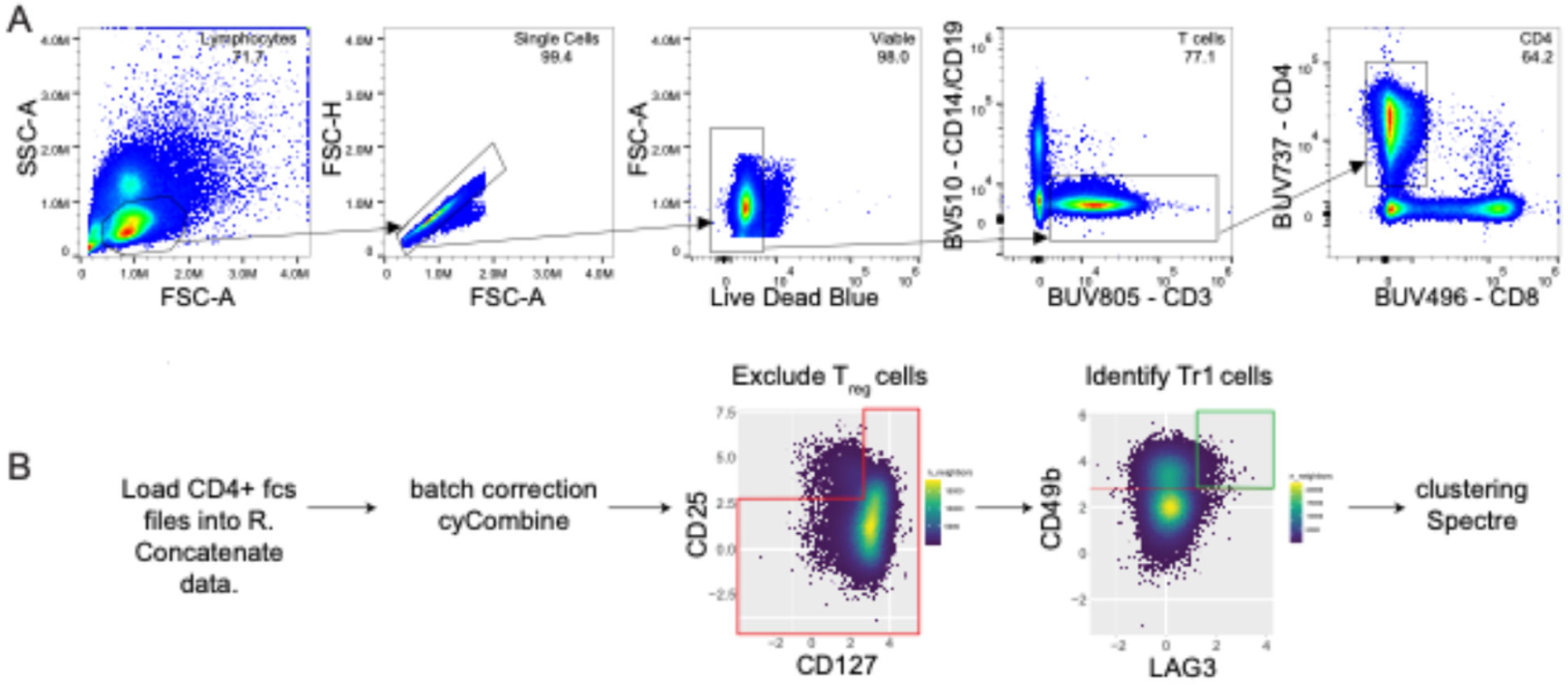
Tr1 cell analysis strategy. **A)** CD4+ T cells were identified based on FSC/SSC, live/dead staining, CD14-CD19-/CD3+, CD4+ cells. **B)** CD4+ cells were exported after gating and analysed in R. Batch correction was performed in cyCombine, Treg cells removed (CD25+CD127low), and Tr1 cells identified based on LAG3+CD49b+. Related to Figure 3.

**Supplementary Figure 6:**
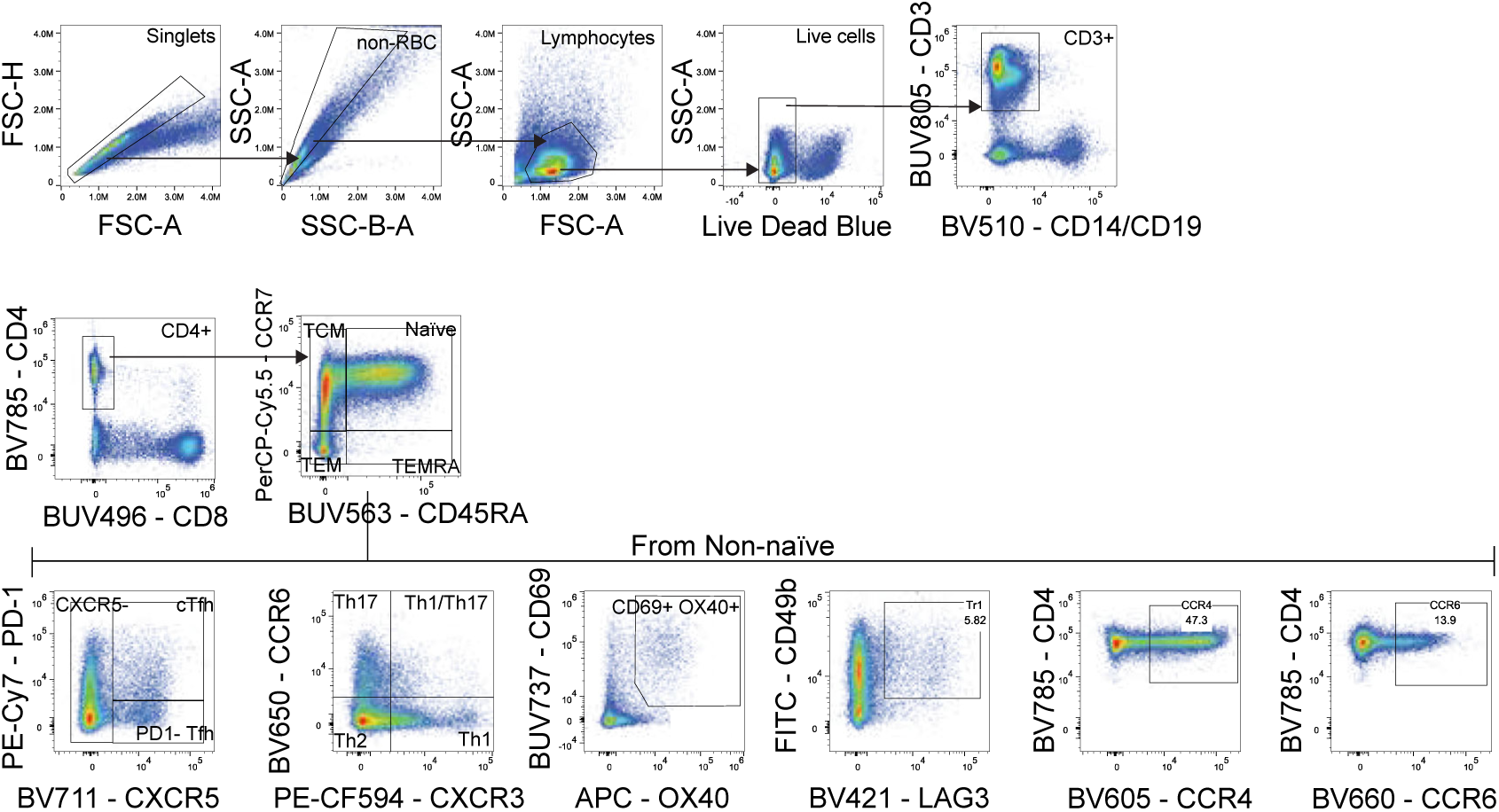
Gating Strategy employed to identify the AIM+ CD4^+^ T cell populations. After gating for the CD45RA^+^ CCR7^+^ naive cells, a “not” gate was used to remove this population and the other CD4^+^ T cell subsets were identified from the non-naive CD4+ T cells. Following this, Boolean gating was using to examine AIM^+^ CD4^+^ T cells. Figure represents PBMCs from Day 15 for D726 in the Placebo group following 18 hour culture with parasitised red blood cells. AND gates were set to analyse all AIM^+^ cells and subset. Related to Figure 4.

**Supplementary Figure 7:**
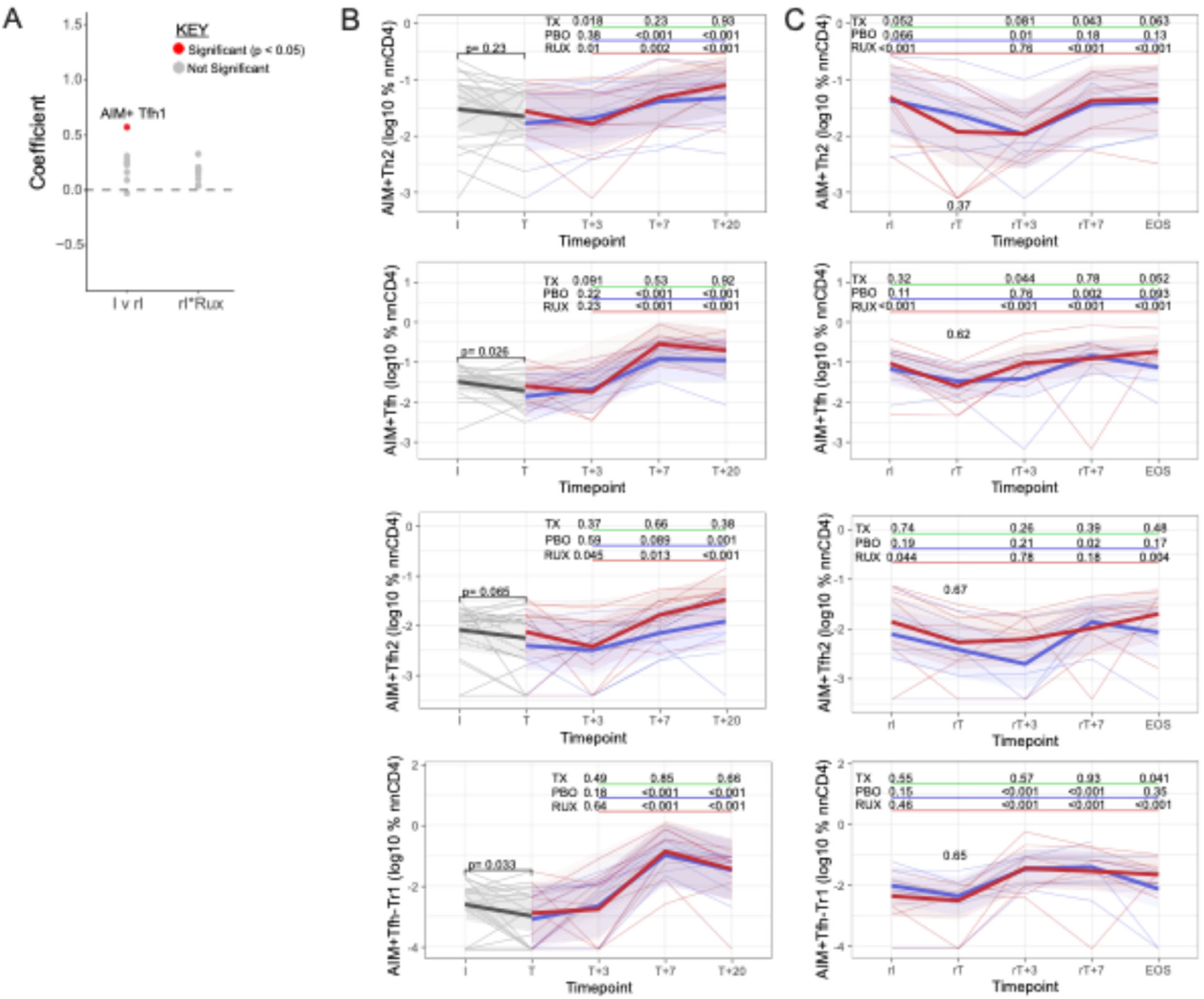
AIM+ CD4 T cell response during first and second infection. Parasite specific CD4 T cells were identified using Activation Induced Marker assay at inoculation (I), treatment (T) and post treatment timepoints (T+3, T+7 and T+20) in first infection, and at re-inoculation (rI), re-treatment (rT) and post treatment timepoints (rT+3, rT+7, EOS) in second infection. **A.** Coefficient of linear mixed models comparing first to second infection inoculation (I verse rI) interaction term of ruxolitinib treatment group. Only AIM+ Tfh1 cells were elevated at rI, and this was not impacted by ruxolitinib. **B/C)** AIM+ Th2, AIM+ Tfh (total Tfh) and AIM+ Tfh2, and AIM+Tfh-Tr1 cells in first and second infection. Data are log10 frequencies of malaria specific AIM+ CD4 T cells, with thin lines representing each participant. Prior to randomisation is in grey, following randomisation ruxolitinib-treated participants are red and placebo are blue. P values are from linear mixed effect models, with bold lines representing the mean of the predicted values from the fitted models for each group. TX is p values for the interaction term between each timepoint (compared to T) and treatment groups (underlined in green). P values for the comparison between each timepoint and T are shown for the placebo (PBO, underlined in blue) and ruxolitinib (RUX, underlined in red) groups, which were determined from contrasts. P value for the difference at rT between placebo and ruxolitinib group, calculated from the intercept is shown at rT.

**Supplementary Figure 8:**
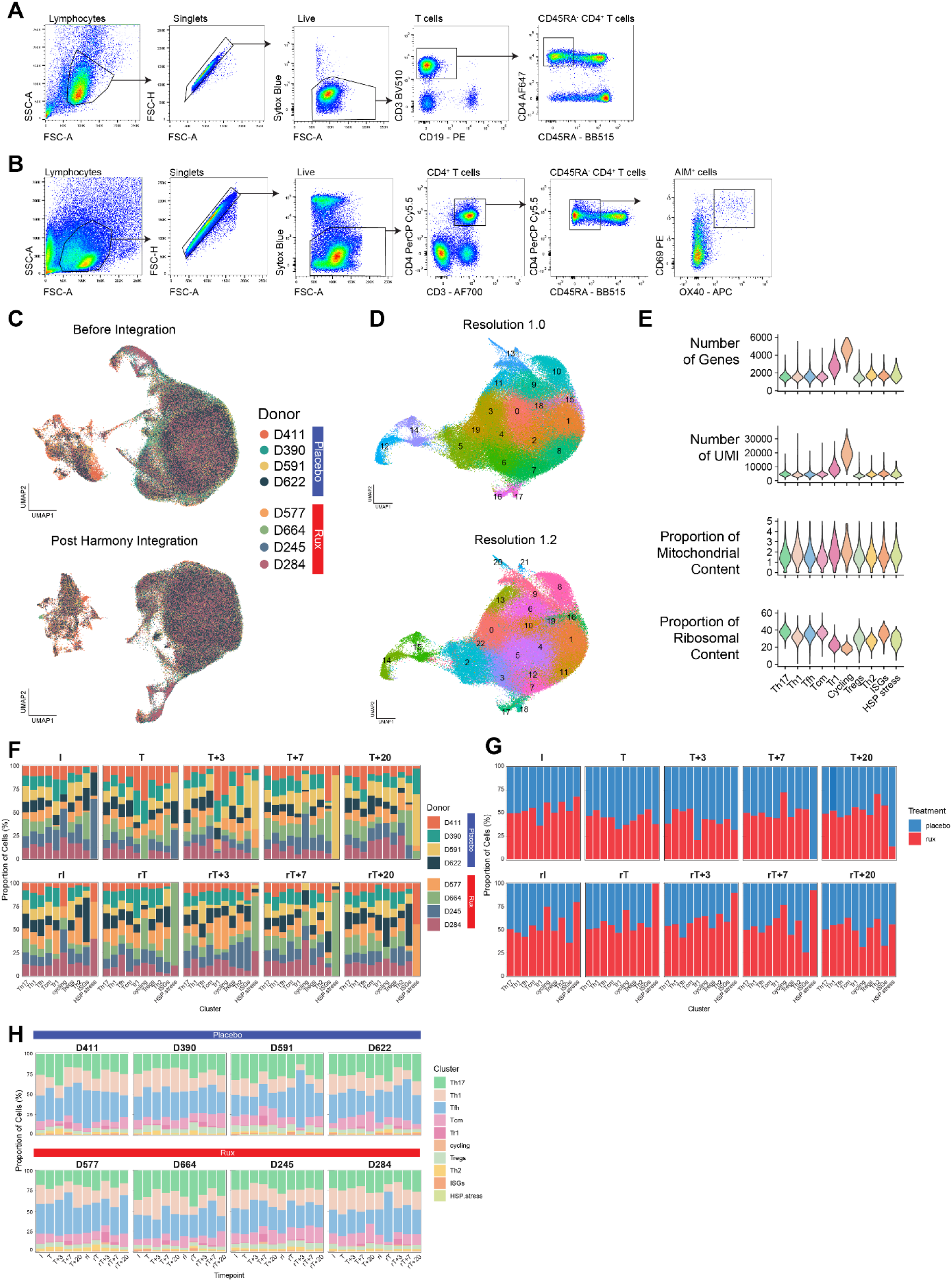
scRNAseq analysis of CD4^+^ T cell responses during first and second infection. **A)** Flow cytometry gating strategy for isolating ex vivo antigen experienced (CD45RA^-^) CD4^+^ T cells and **B)** AIM^+^ (CD69^+^OX40^+^) CD4^+^ T cells. **C)** UMAP visualisation of ex vivo and AIM^+^ CD4^+^ T cells coloured by donor prior to and post-Harmony integration by donor. **D)** UMAP visualisation showing clustering of the ex vivo CD4^+^ T cells at resolution 1 and 1.2. **E)** Violin plots showing quality control metrics for the ex vivo CD4^+^ T cells. **F)** Stacked bar graphs split by timepoint showing the proportion of donor or **G)** treatment group per ex vivo CD4^+^ T cell cluster. **H)** Stacked bar graphs split by donor showing the proportion of ex vivo CD4+ T cell cluster per timepoint.

**Supplementary Figure 9:**
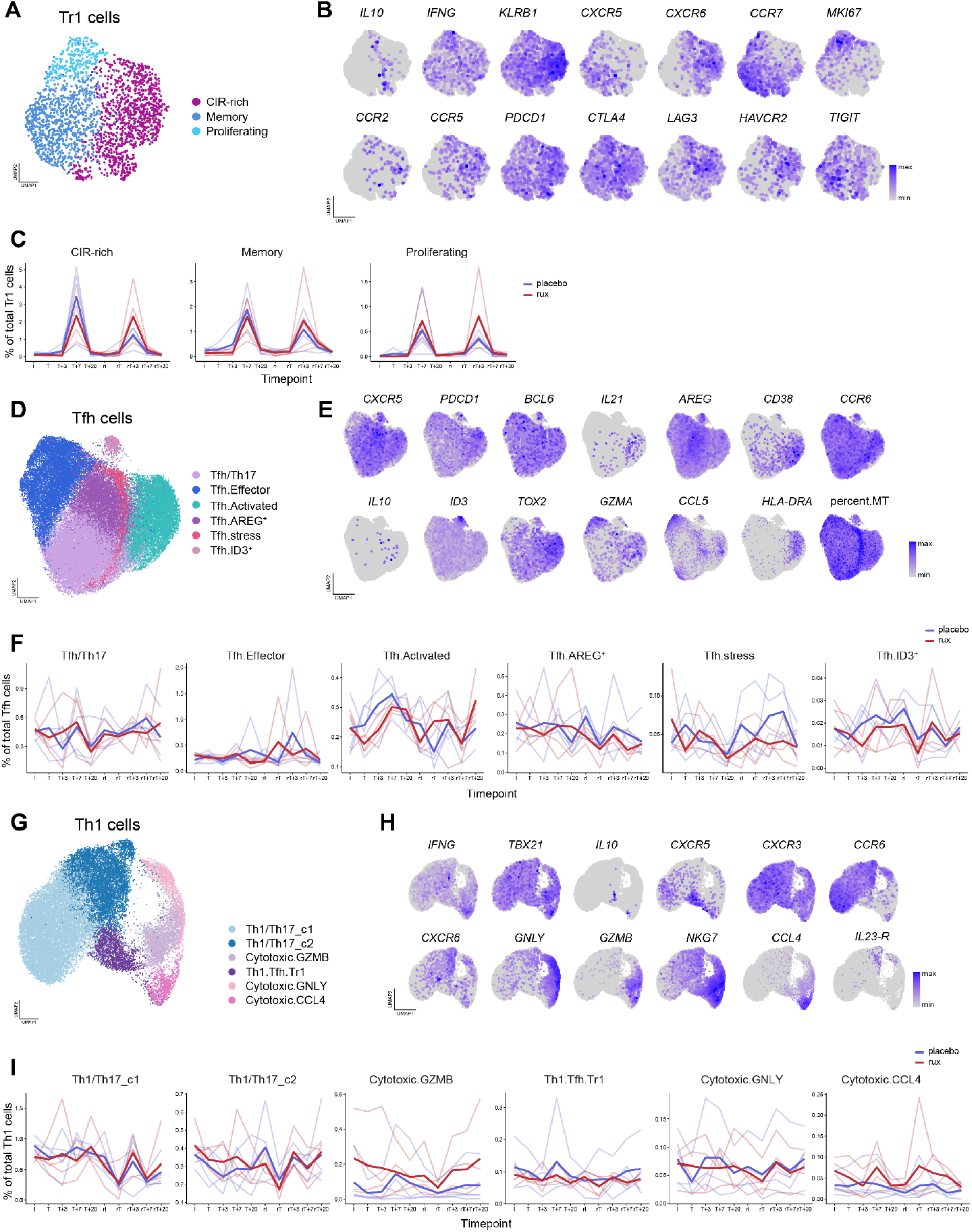
scRNAseq Th1-Tfh-Tr1 axis cells re-clustering. **A)** UMAP visualisation shows three clusters of Tr1 cells after subsetting and re-clustering and **B)** expression of select cluster marker genes. **C)** Line graphs showing the cluster frequency as a percentage of total Tr1 cells during first and second infections. Lines are coloured by treatment group, with bold lines representing group means and transparent lines showing frequencies for each participant. **D)** UMAP visualisation shows six clusters of Tfh cells after subsetting and re-clustering and **E)** expression of select cluster marker genes. **F)** Line graphs showing the cluster frequency as a percentage of total Tfh cells during first and second infections. Lines are coloured by treatment group, with bold lines representing group means and transparent lines showing frequencies for each participant. **G)** UMAP visualisation shows six clusters of Th1 cells after subsetting and re-clustering and **H)** expression of select cluster marker genes. **I)** Line graphs showing the cluster frequency as a percentage of total Th1 cells during first and second infections. Lines are coloured by treatment group, with bold lines representing group means and transparent lines showing frequencies for each participant.

**Supplementary Figure 10:**
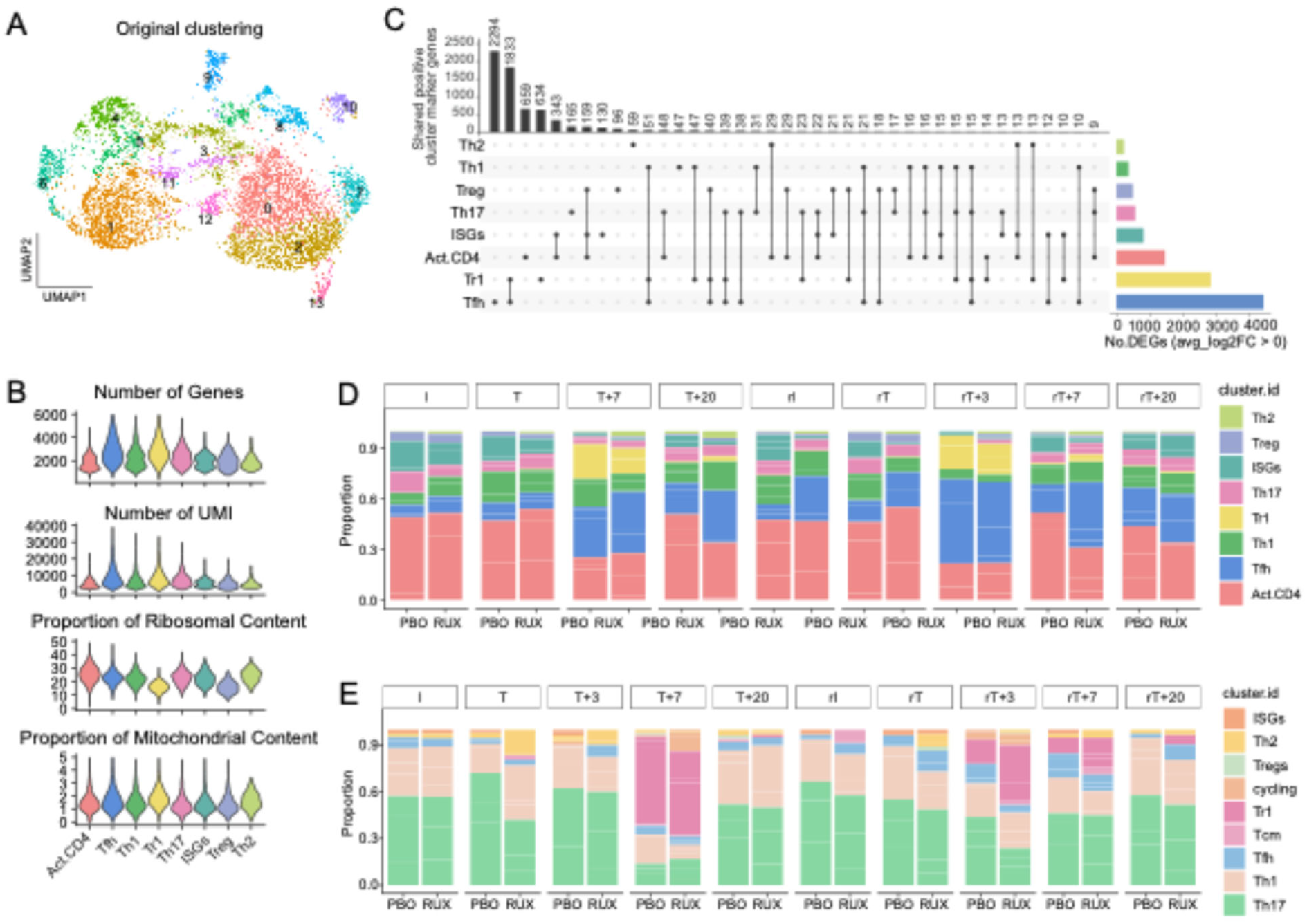
AIM+ scRNAseq data and malaria specific cells. **A)** UMAP visualisation showing clustering of the AIM+ CD4^+^ T cells. **B)** Violin plots showing quality control metrics for the AIM+ CD4^+^ T cells. **C)** Upset plot showing the number of shared marker genes between clusters in AIM+ data set. The bar graph on the right shows the total number of differentially expressed cluster marker genes for each subset. **D)** Subset proportions of AIM+ CD4 T cells at each timepoint, by treatment group. **E)** Subset proportions of malaria-specific cells identified within ex vivo data set based on TCR analysis of AIM+ data set.

